# CACNA1B gain-of-function mutation drives familial atrioventricular nodal re-entrant tachycardia through sympathetic hyperactivity

**DOI:** 10.1101/2022.03.09.22271906

**Authors:** Rong Luo, Xin Cao, Chenqing Zheng, Yuanlin Tang, Qianzi Tang, Qin Sheng, Hongmei Deng, Lei Yao, Zhixue Wang, Wei-Wen Lim, Xiushan Wu, Xiaoping Li

## Abstract

**Background:** Familial clustering of atrioventricular nodal re-entrant tachycardia (AVNRT) suggests a genetic basis, yet the underlying molecular etiology remains undefined. *CACNA1B* encodes the α1 subunit of the N-type calcium channel (Ca_V_2.2), a key regulator of sympathetic neurotransmission. We identify *CACNA1B* as a novel causative gene for familial AVNRT and define its neurogenic mechanism of arrhythmogenesis.

**Methods:** Whole-exome sequencing of a four-generation pedigree with autosomal dominant AVNRT revealed a novel heterozygous missense variant, *CACNA1B* p.K567R. Functional effects were assessed in zebrafish and an orthologous *Cacna1b* p.K565R knock-in rat using ambulatory telemetry, programmed electrical stimulation, patch-clamp electrophysiology, ultrastructural analyses, and integrated multi-omics of superior cervical ganglion (SCG) and AV nodal tissues. Pharmacological interventions were conducted using the selective N-type calcium channel inhibitor, Ω -conotoxin GVIA, and the clinically approved L- and N-type calcium channel blocker, cilnidipine.

**Results:** The *CACNA1B* p.K567R variant co-segregated with AVNRT and induced an accelerated cardiac rhythm in the mutant overexpressed zebrafish. Patch-clamp analysis demonstrated a marked Ca_V_2.2 gain-of-function (GoF) driven by increased surface channel density. *Cacna1b* p.565R knock-in rats exhibited chronic sympathetic overactivity with significantly perturbed heart rate variability (HRV), dual AV nodal pathways with atrial echo beats, and inducible AVNRT. Single-cell and multi-omics profiling in SCG revealed functional hyperactivation of the glutamatergic subpopulation and upregulation of presynaptic exocytotic components (SYT1/SNAP25), producing sustained sympathetic hyperexcitability. This neurogenic drive triggered metabolic reprogramming and oxidative stress in downstream AV nodal transitional cells, establishing an arrhythmogenic substrate for reentry. Treatment with ω-conotoxin GVIA and cilnidipine reversed the hyperadrenergic phenotype and normalized cardiac autonomic tone and heart rate.

**Conclusions:** *CACNA1B* is a novel disease gene of familial AVNRT. Ca_V_2.2 GoF induces sympathetic hyperactivity and secondary AV nodal remodelling, establishing a neurogenic substrate for reentrant tachycardia, and N-type calcium channel blockade represents a promising precision therapeutic strategy for *CACNA1B*-mediated, autonomic-driven arrhythmias.

**What is known:**

- Familial clusters of AVNRT suggest genetic predisposition consistent with autosomal dominant inheritance but mechanisms unknown.
- Sympathetic tone is a critical modulator of AV nodal conduction and a primary determinant of AVNRT susceptibility.

**What new information does this article contribute?:**

- *CACNA1B* (p.K567R) gain of function variant overexpression in HEK293T cells and zebrafish embryos demonstrate heart rate and electrophysiological abnormalities implicates *CACNA1B* as a disease gene of familial AVNRT.
- A *Cacna1b* (p.K565R) orthologous rat model recaptures sympathetic hyperactivity, severe autonomic dysregulation and arrhythmia susceptibility.
- Ca_V_2.2 hyperactivity drives chronic sympathetic overactivation through SYT1-SNAP25 and secondary AV nodal remodeling.

**Novelty and Significance:** This work reframes the pathogenesis of familial AVNRT by establishing *CACNA1B* gain of function as a primary neurogenic driver of disease. Rather than arising solely from an anatomic dual pathway circuit, AVNRT can originate from genetically driven sympathetic hyperexcitability that secondarily remodels the AV node. The discovery that Ca_V_2.2 overactivity expands glutamatergic sympathetic neurons, enhances presynaptic exocytotic cargo, and chronically hyperactivates the AV node provides a mechanistic bridge between autonomic dysfunction and reentrant arrhythmia. Cilnidipine, a clinically available dual L-/N type calcium channel blocker, normalizes sympathetic overactivity. These findings portray *CACNA1B* as a potential target and position N type calcium channel inhibition as a mechanism based therapeutic strategy for familial or refractory AVNRT. The study broadens the conceptual framework of AVNRT, highlights the autonomic nervous system as a modifiable arrhythmogenic source, and provides a translatable treatment avenue for patients carrying *CACNA1B* variants.

## Introduction

Atrioventricular nodal re entrant tachycardia (AVNRT) is the most common paroxysmal supraventricular tachycardia and classically arises from longitudinal dissociation within the AV (atrioventricular) node as dual fast and slow AV nodal pathways to sustain re-entry.^1^ Although catheter ablation is effective, the molecular determinants governing AV nodal conduction remains poorly defined.^2,3^ Familial clustering strongly suggests a genetic etiology,^4–6^ yet unlike *PRKAG2*-mediated Wolff-Parkinson-White syndrome, no definitive causative gene has been identified.^7,8^

Recent whole-exome sequencing (WES) studies have implicated rare variants in cardiac ion-handling genes,^9,10^ and our prior work showed enrichment of ion channel variants in sporadic AVNRT,^11^ highlighting calcium signaling as a potential pathogenic pathway.^12^ However, these associations lacked mechanistic resolution and did not explain how specific channel defects generate the functional substrate required for AV nodal reentry. Thus, the molecular aetiology of familial AVNRT remains unresolved.

*CACNA1B*, encoding the α1B subunit of the N-type voltage-gated calcium channel (Ca_V_2.2), is a key regulator of presynaptic Ca² influx and neurotransmitter release in sympathetic neurons. Unlike myocardial L-type channels that mediate excitation–contraction coupling,^13^ Ca_V_2.2 directly interfaces with the SNARE complex to regulate neurotransmitter release.^14,15^ Given the AV node’s role as an autonomic integration hub, perturbations in Ca_V_2.2-dependent neurotransmission could destabilize nodal conduction. Sympathetic activation is known to shortens AV nodal refractoriness and promotes AVNRT. This concept aligns with clinical observations that catecholaminergic stress facilitates AVNRT initiation and maintenance.^16^

Here, we identify a pathogenic *CACNA1B* gain-of-function (GoF) variant (p.K567R) in a large pedigree with autosomal dominant AVNRT. Using an orthologous knock-in rat model and systems-level multi-omics, we define a novel neurogenic mechanism in which intrinsic sympathetic hyperactivity remodels the AV nodal substrate to permit reentry. Finally, we demonstrate that L-/N-type calcium channel inhibition with cilnidipine restores impaired heart rate variability and attenuates the elevated heart rate, establishing a mechanism-based therapeutic strategy for *CACNA1B*-related AVNRT.

## Methods

### Data availability

All publicly available datasets are deposited in the repositories listed below. The authors declare that all supporting data are available within the article and in the Data Supplement. Due to the sensitive nature of patient information, de-identified clinical data requests can be made to the corresponding author upon reasonable request. A Major Resources Table is included in the Data Supplement (**Supplementary Table S1**).

### Ethics approval

Human studies were approved by the Institutional Review Board of Sichuan Provincial People’s Hospital (ID #2021 239); all participants provided written informed consent. All animal procedures were approved by the Institutional Animal Care and Use Committee of Chengdu Medical College (ID #2022 423).

### Patient cohort

A four-generation pedigree with autosomal dominant AVNRT was recruited from our established cohort.^12^ Clinical evaluation included electrocardiography (ECG), Holter monitoring, and echocardiography; diagnoses were confirmed by invasive electrophysiology or transesophageal pacing. A validation cohort of 86 sporadic AVNRT cases was screened for variant prevalence.^11^ WES was performed on four affected family members, and variants were filtered by rarity (MAF <0.001), conservation, and predicted deleteriousness. Candidate variants were validated by Sanger sequencing, and assessed for co-segregation in nine relatives.

### Zebrafish overexpression assay

Functional testing of *CACNA1B* p.K567R was performed by Nanjing YSY Biotech Company. Capped mRNAs encoding human wild-type (WT) or mutant *CACNA1B* were synthesized using SP6 after NotI linearization (MEGAclear Kit, Thermo Fisher). One-cell AB-strain embryos were microinjected with 1 nL mRNA (50-200 ng/µL). Embryos were maintained at 28.5°C under a 14-/10-h light-dark cycle. Gross morphology and cardiac rhythmicity were assessed at 48, 72, and 120 hours post-fertilisation (hpf). Heart rate was quantified in anesthetized embryos (0.2 mg/mL tricaine; n=20/group) by manual ventricular beat counts over 20 seconds. No dose-dependent teratogenicity was observed.

### RNA sequencing

RNA libraries were prepared using NEBNext® Ultra™ Kit and sequenced on an Illumina NovaSeq 6000. Reads were aligned with HISAT2, quantified by featureCounts, and analyzed using DESeq2 (adjusted P<0.05). Functional enrichment was performed with clusterProfiler.

### Generation of *Cacna1b* mutant rats

An orthologous *Cacna1b* p.K565R knock-in rat was generated on a Sprague-Dawley background using CRISPR/Cas9 (GemPharmatech, Nanjing, China). A single-nucleotide substitution (c.1694A>G) introduced into exon 12 produces the p.K565R missense change corresponding to human p.K567R. Genotypes were confirmed by PCR and Sanger sequencing (**Supplementary Figure 1A–C**). WT (*Cacna1b*^+/+^), heterozygous (*Cacna1b*^+/K565R^, HET) and homozygous (*Cacna1b*^K565R/K565R^, HOM) littermates (8-12 weeks; 200–300 g) of both sexes were used. Rats were housed under specific pathogen-free (SPF) conditions (23±1°C; 12/12-h light-dark cycle) with ad libitum food and water. Potential CRISPR/Cas9 off-target sites were predicted in silico, and the top 10 candidate loci were examined by PCR and Sanger sequencing (Supplementary Table 1), with no detectable off-target editing identified (data not shown).

### Electrocardiography and telemetry

Rats were anesthetized with pentobarbital sodium (40 mg/kg) and implanted with ECG transmitters (Data Sciences International, DSI, United States) as described previously.^17^ After a two-week recovery, continuous ECGs were recorded in freely moving animals. Heart rate variability (HRV) was analyzed with Ponemah software (DSI). Time-domain indices (standard deviation of NN intervals, SDNN; root mean square of successive differences, RMSSD) quantified overall and parasympathetic variability; frequency-domain low-frequency (LF) and high-frequency (HF) power yielded the LF/HF ratio as an index of sympathetic activity.^18,19^ Poincaré plots were generated during the active phase (16:00 hours) for non-linear heart rate dynamics.

### Baroreflex sensitivity (BRS)

After hemodynamic stabilization, BRS was assessed using a phenylephrine pressor challenge (200 µg/kg, i.v.) to elicit a rise in systolic blood pressure (SBP). Beat-to-beat SBP and RR intervals were recorded continuously. BRS index was calculated as the slope of RR-SBP regression (ms/mmHg) during the ascending phase of the pressor response. Sympathetic nerve activity was recorded concurrently during vasoactive challenge with bipolar platinum electrodes and quantified using spike histogram analysis in LabChart. Discharge frequency before and after phenylephrine challenge was quantified to assess reflex sympathetic suppression.

### Pharmacological interventions

Acute autonomic challenges were performed in conscious rats with established telemetry baselines. Sympathetic and parasympathetic influences were assessed using the selective β1-adrenergic blocker metoprolol (5 mg/kg, i.p.) and the muscarinic antagonist atropine (1 mg/kg, i.p.), respectively.^20^ Sympathetic reserve was tested with the non-selective β-agonist isoprenaline (0.5 mg/kg, i.p.). N-type channel dependence was evaluated using omega (Ω)-conotoxin GVIA (10 µg/kg, i.p.).^21^ For therapeutic rescue, rats received cilnidipine (10 mg/kg/day, p.o.) for 3 weeks, a dual N/L-type calcium channel blocker known to suppress sympathetic neurotransmission.^22^ Telemetric HR and HRV indices were compared pre- and post-treatment.

### Intracardiac electrophysiology

Rats were anesthetized with ethyl carbamate, intubated, and mechanically ventilated as previously described.^17,23^ A 3F octapolar electrode catheter (SMC-304, Physio-Tech, Germany) was advanced via the right jugular vein for intracardiac recording and pacing. Programmed electrical stimulation (PES) was delivered at twice the diastolic threshold using 3-ms rectangular pulses (SEN-8203; Jingjiang Inc., China). Atrial, atrioventricular and ventricular effective refractory periods (AERP, AVNERP, and VERP respectively) were measured at basic cycle lengths (BCL) of 150, 120, and 100 ms using an S1(×8)-S2 protocol. ERP was defined as the longest S1-S2 interval failing 1:1 capture. Wenckebach cycle length (WCL) was determined by decrementing pacing cycle length by 1-ms after every four stimuli until atrioventricular block (AVB) occurred. Arrhythmias were classified per Lambeth Conventions.^24^ Dual AV nodal physiology (DAVNP) was defined by an atrial-His (AH) interval “jump” of ≥10 ms in response to a 1–2 ms decrement in S1–S2 coupling interval, indicating fast-to-slow pathway shift. Echo beats or AVNRT during premature or burst atrial pacing confirmed DAVNP.^25,26^

### Heterologous expression of the Ca_V_2.2 minimal functional complex

Human Ca_V_2.2 channels were expressed in human embryonic kidney 293T (HEK293T) cells maintained in Dulbecco’s modified eagle medium with 10% fetal bovine serum and 1% penicillin-streptomycin. Cells were transient transfected with plasmids encoding WT or p.K567R *CACNA1B* α1B and the cytosolic CACNB3 auxiliary β3 subunit (1:1 ratio) using Lipofectamine 3000 (Invitrogen) according to the manufacturer’s instructions. A minimal α1B+β3 configuration was used to isolate α1-subunit-specific biophysical effects.^27^

### Isolation of superior cervical ganglia (SCG) neurons

SCG neurons from 8–12-week-old WT, HET, and HOM rats were desheathed, minced, and enzymatically digested (collagenase type II 1.5 mg/mL; trypsin 0.25%) for 45–60 min at 37°C. After trituration and centrifugation (800 rpm for 3 minutes), cells were resuspended in Neurobasal-A medium supplemented with 2% B27, 2 mM L-glutamine, and 50 ng/mL nerve growth factor (NGF; Alomone Labs) plated on poly-L-lysine/laminin (100 µg/mL, 20 µg/mL; respectively)-coated coverslips, and electrophysiological recorded within 24–36 hours to minimize culture-induced changes.^28^

For primary cultures, sympathetic neurons were isolated from 1-to-3-week-old Sprague-Dawley rats. After urethane anesthesia (0.5-1.0 mL/100 g), the bilateral SCG were excised into ice-cold Ca² -free Tyrode’s solution (pH 7.4), decapsulated, minced and digested with Liberase and DNase for 30 minutes at 37°C. Digestion was halted with ice-cold, NGF-free Neurobasal medium washed twice. Tissues were gently triturated, centrifuged, resuspended in complete Neurobasal medium and plated onto coverslips (as above), and incubated at 37°C for 1-2 hours to facilitate cellular adhesion before imaging.

### Cellular electrophysiology

Whole-cell patch clamp recordings on HEK293T and rat SCG neurons were performed by Beijing Haiwei Panstone Biotechnology Co., Ltd. HEK293T cells (24–48 h post-transfection) and SCG neurons were recorded at 22–24°C using MultiClamp 700B amplifiers. Signals were digitized at 10 kHz and filtered at 2 and 2.9 kHz (SCG and HEK293T respectively). Pipettes (2–5 MΩ) contained a cesium-based internal solution (in mM): 140 CsCl, 20 TEA-Cl, 10 HEPES, 0.1 BAPTA, 1 MgCl, 0.1 leupeptin, 4 Mg-ATP, 0.3 Na-GTP (pH 7.3, adjusted with CsOH). The external solution contained 10 mM Ba² as charge carrier (in mM): 140 TEA-Cl, 10 BaCl, 1 MgCl, 10 HEPES, and 10 glucose (pH adjust 7.4 with TEA-OH). SCG recordings included 1 µM Tetrodotoxin (TTX) to suppress voltage-gated Na spikes.^29^ Series resistance was compensated by 70–80% and cells were held at −90 mV. Depolarizing steps (200-ms) were applied from −30 to +60 mV (HEK293T) or −30 to +40 mV (SCG) in 5-mV increments. Peak current density (pA/pF) was normalized to capacitance (Cm). Conductance-voltage (G–V) curves were fitted with Boltzmann functions to derive steady-state activation kinetics (V_1/2_ and k).^30^

### SCG and AV node snRNAseq

To map the neuro-cardiac regulatory network, SCG and AV node tissues from WT and mutant rats were processed using the 10x Genomics platform for single-nucleus RNA sequencing. Reads were aligned and quantified using CellRanger; downstream analyses were performed with Seurat.^31^ Batch effects were corrected using Harmony,^32^ and mutation-associated gene modules were identified using high-dimensional Weighted Gene Co-expression Network Analysis (hdWGCNA).^33^

### Liquid chromatography tandem mass spectrometry (LC-MS/MS)

SCG samples were digested with trypsin and labeled using TMTpro 16-plex reagents. Fractionated peptides (Dionex UltiMate-3000) were analyzed by LC-MS/MS (Orbitrap Exploris 480). Data were processed with MaxQuant (v1.6.6) using the *Rattus norvegicus* UniProt database (1% FDR). Search parameters included standard TMT modifications, trypsin specificity, and a mass tolerance of 20 ppm. Bioinformatics analyses for Gene Ontology (GO) enrichment and Kyoto Encyclopedia of Genes and Genomes (KEGG) pathways were conducted using DAVID.^34^

### Metabolomics analysis

Metabolites were extracted in methanol containing L-2-chlorophenylalanine and analyzed using an Agilent 1290 UHPLC system coupled to an AB Sciex Triple TOF 5600 mass spectrometer. Data were processed using XCMS,^35^ and metabolites were annotated using HMDB,^36^ and KEGG.^37^ Pathway enrichment was performed using Metascape and MetaboAnalyst.

### Histology and immunofluorescence

SCG neurons were fixed with 4% paraformaldehyde, permeabilized, and incubated with primary and FITC-conjugated secondary antibodies (**Supplementary Table 2**). Images were acquired using a Nikon A1R confocal microscope. Fluorescence intensity was quantified by ImageJ.^38^

### Transmission electron microscopy (TEM)

Myocardial blocks (1 mm³) were fixed in 2.5% glutaraldehyde and 1% osmium tetroxide, underwent graded dehydration, embedded in Epon 812, sectioned (70-90 nm), and stained with uranyl acetate and lead citrate. Images were acquired using a HITACHI HT7800 (80 kV) to assess mitochondrial and structural morphology.

### Molecular and biochemical analysis

SCG tissue was processed for RNA and protein extraction. RT-qPCR was normalized to β*-actin* expression. Protein lysates were analyzed by SDS-PAGE and immunoblotting. Primer sequences for RT-qPCR and antibodies are provided in **Supplementary Table 2**.

### Statistical analysis

Data are presented as mean ± SD unless otherwise stated. Normality was assessed using the Shapiro-Wilk test. Two-group comparisons used unpaired or Welch’s t-test as appropriate; categorical variables used Fisher’s exact test. Hierarchical cellular data were analyzed using linear mixed-effects models with animal ID as a random effect to avoid pseudoreplication.^39^ Analyses were performed using SPSS 26.0 and GraphPad Prism 6.0. Statistical significance was established at P<0.05.

## Results

### Identification of CACNA1B p.K567R in familial AVNRT

We evaluated a four-generation pedigree exhibiting autosomal dominant arrhythmias (Figure 1A-D). Affected members showed variable expressivity. The proband reported paroxysmal palpitations with abrupt onset and termination. Twelve-lead ECG documented paroxysmal supraventricular tachycardia (PSVT), and echocardiography showed normal cardiac structure and function (LVEF 68%). Although baseline Holter recordings demonstrated sinus rhythm, HRV indices were reduced (SDNN 96 ms, SDANN 82 ms, rMSSD 24 ms, pNN50 3%) indicating autonomic impairment. Subsequent electrophysiological study (EPS) confirmed typical slow-fast AVNRT, successfully treated by radiofrequency catheter ablation (RFCA).

**Figure 1.**
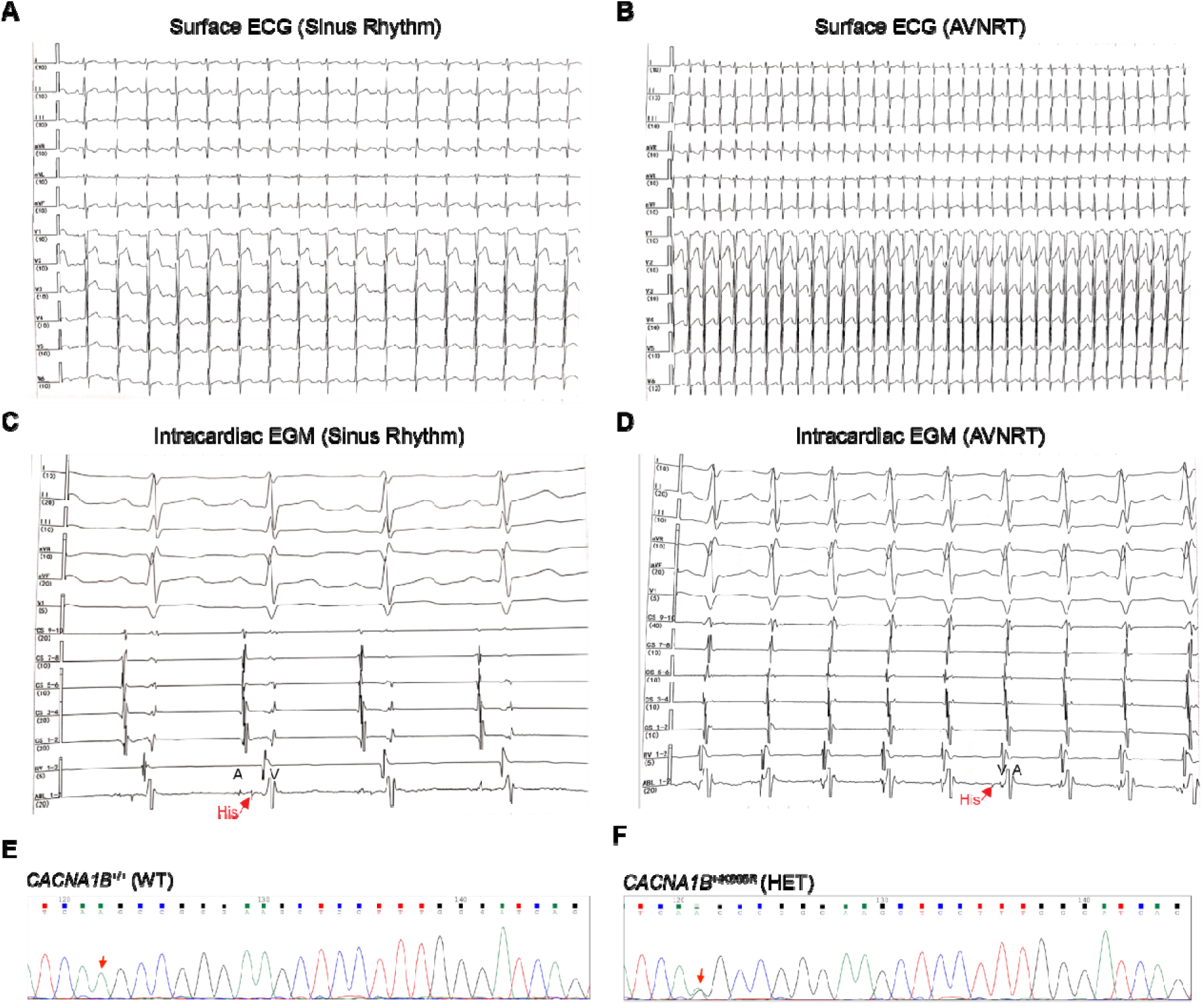
Clinical presentation and genetic identification of the familial AVNRT pedigree carrying the *CACNA1B* p.K567R mutation. **(A)** Representative 12-lead surface ECG of the proband during normal sinus rhythm, exhibiting unremarkable P-wave morphology and PR intervals. **(B)** 12-lead surface ECG captured during a symptomatic episode, documenting regular, narrow-complex supraventricular tachycardia (SVT). Retrograde P waves are visibly buried within the terminal portion of the QRS complex (presenting as a characteristic pseudo-r’ wave in lead V1). **(C)** Baseline intracardiac electrograms (EGM) during sinus rhythm, demonstrating intact anterograde conduction with normal AH and HV intervals. His is indicated by the red arrow with atrial (A) and (ventricular (V) activation. **(D)** Intracardiac EGM during the tachycardia episode. The simultaneous A and V activation (A on V) with a short VA interval confirms the diagnosis of typical slow-fast AVNRT. **(E-F)** Representative Sanger DNA sequencing chromatograms. The panel identifies the heterozygous missense mutation c.1700A>G (p.K567R) in the *CACNA1B* gene of the affected members (F), contrasted with the wild-type (WT) sequence in healthy familial members (E).

One of her parents displayed a similar phenotype, with Holter monitoring revealing paroxysmal sinus tachycardia and typical AVNRTA despite the echocardiogram being unremarkable. RFCA achieved complete resolution. One of the proband’s siblings was diagnosed with typical AVNRT by transesophageal atrial pacing and managed conservatively due to infrequent episodes. In contrast, one of the proband’s relatives had AVRT mediated by a left lateral accessory pathway, also cured by RFCA.

WES of three affected individuals and one unaffected parent identified a heterozygous *CACNA1B* missense variant, c.1700A>G (p.K567R), located in the voltage-sensing region of the conserved Ca_V_2.2 α1B subunit DII–S2 transmembrane segment. Pathogenicity predictions were uniformly high (REVEL 0.872; GERP++ RS 4.68; phyloP470way 9.23), fulfilling ACMG PP3_supporting criterion. Sanger sequencing demonstrated complete co-segregation across three generations (PP1_moderate) and absence from population databases supporting rarity (PM2_supporting). Together, these criteria (PP1_moderate, PM2_supporting, PP3_supporting) classifies p.K567R as likely pathogenic according to the ACMG/AMP guidelines.^40^ Screening of 86 sporadic AVNRT cases^11^ identified three additional rare *CACNA1B* missense variants (3/86, 3.5%): c.493G>A (p.G165S) in one patient and c.6744G>C (p.Q2248H) in two others, suggesting broader relevance to both familial and sporadic AVNRT (**Supplementary Table S2**).

### Zebrafish *CACNA1B* p.K567R overexpression causes chronotropic GoF

Transient overexpression of human *CACNA1B* mutant mRNA in zebrafish embryos produced a clear gain-of-function phenotype. Embryos expressing p.K567R exhibited significantly elevated heart rates at 48 hpf (168.0±1.5 vs. 147.3±2.8 bpm; p<0.01) and 72 hpf (177.4±4.6 vs. 156.8±2.3 bpm; p<0.01), with low teratogenicity (<6%) (**Figure 2A-B**). Whole-embryo transcriptomics showed early enrichment of “Adrenergic signaling in cardiomyocytes” and “Calcium signaling pathway” at 48 hpf, persisting at 72 hpf with additional activation of “Cardiac muscle contraction” (**Figure 2C-E**).^37^ Downregulated pathways involved glycolysis and ribosome biogenesis, suggesting an early bioenergetic trade-off driven by this hyperactive state. These data indicate early enhancement of sympathetic and Ca² handling programs, consistent with increased arrhythmia susceptibility.^41,42^

**Figure 2.**
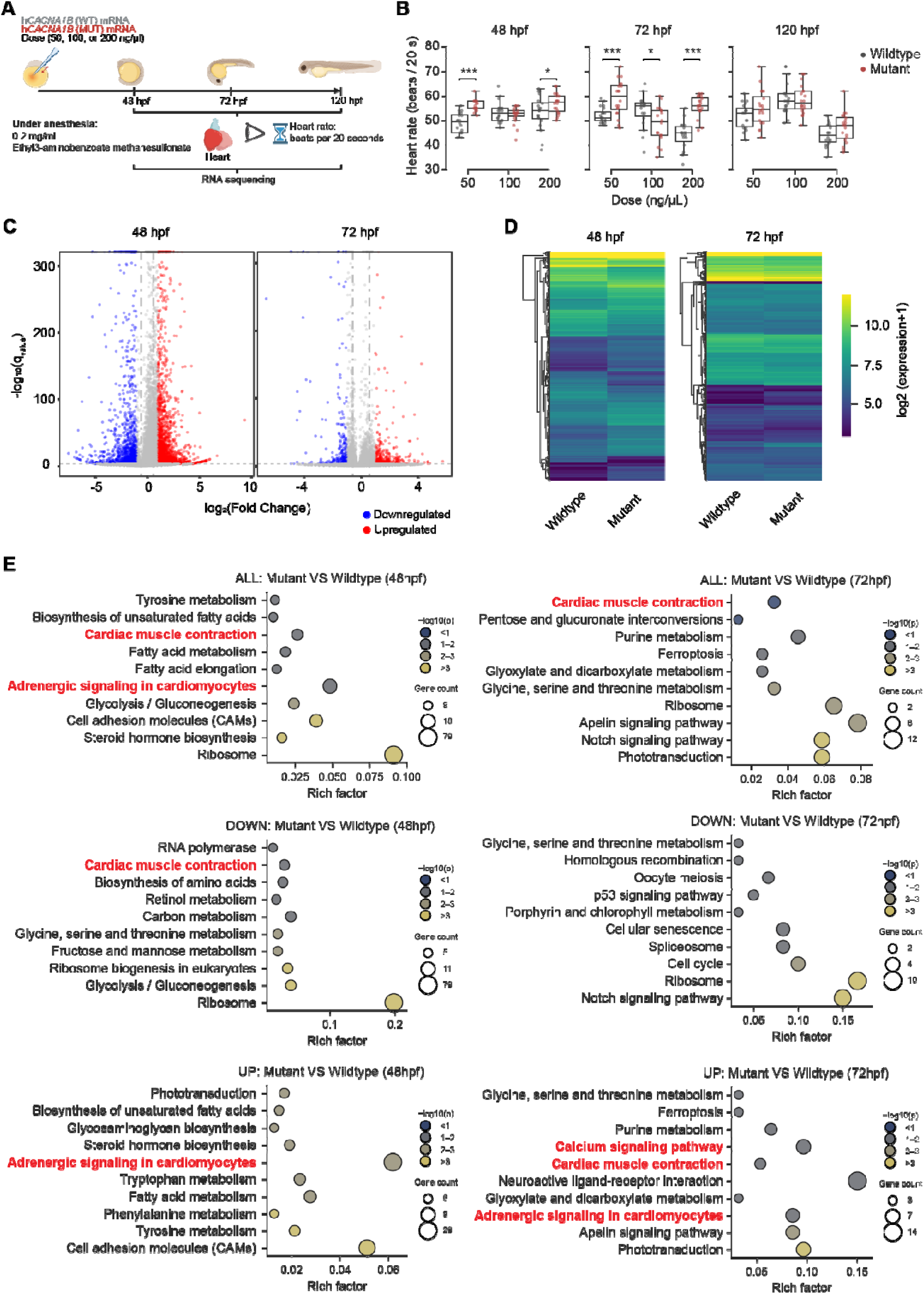
*In vivo* validation reveals a chronotropic gain-of-function (GoF) and a hyperadrenergic transcriptomic signature in mutant zebrafish. **(A)** Schematic of experimental workflow of zebrafish embryos microinjected with wildtype (WT) or mutant (MUT) human *CACNA1B* (*hCACNA1B*) mRNA at doses of 50, 100, or 200 ng/μl. Created with BioRender.com. **(B)** Quantification of atrial heart rate (beats per 20 seconds) in zebrafish embryos evaluated at 48, 72, and 120 hours post-fertilization (hpf) confirming positive chronotropic effects. Data are presented as median ± IQR (n=20 embryos/group; *P < 0.05, ***P < 0.001). Statistical analyses by student t-test. **(C-E)** Volcano plots **(C)**, expression heatmaps **(D)**, and bubble plots **(E)** of differentially expressed genes (DEGs) between WT and MUT zebrafish samples at 48 and 72 hpf. DEGs were defined using FDR < [0.001] and |log2FC| ≥ [1]. Bubble plots illustrating functional KEGG pathway enrichment analyses of all (top), downregulated (mid) and upregulated (bottom) DEGs. Transcriptomic profiling reveals a specific and significant enrichment of pathways associated with “Adrenergic signaling in cardiomyocytes,” “Calcium signaling pathway,” and “Cardiac muscle contraction” in the MUT group.

### Normal structural developmental but evidence of neurogenic sympathetic activation

To mimic the human mutation *in vivo*, we generated *Cacna1b* p.K565R knock in rats (**Supplementary Figure 1**). WT, HET, and HOM animals exhibited normal growth, morphology, and echocardiographic parameters (**Supplementary Table S3**). Despite this, mutants exhibited elevated heart rate and increased systolic and mean arterial pressures, with preserved diastolic pressure (**Supplementary Figure 2A-C**).

Because Ca_V_2.2 channels regulate neurotransmitter release at sympathetic terminals, we quantified circulating catecholamines and their metabolites. Levels of rapidly degraded catecholamines, norepinephrine (NE), dopamine (DA), and epinephrine (E), were similar across genotypes. The stable norepinephrine (NE) metabolite normetanephrine (NMN) trended higher in mutants (*P*=0.078), while the metabolite of dopamine (DA) 3-Methoxytyramine (3-MT) and epinephrine (E) metabolite metanephrine (MN) remained unchanged (**Supplementary Figure 2D-F**). This selective NMN elevation indicates chronic sympathetic nerve-terminal activation, without adrenal output.^43^

### Sympathetic hyperactivity and autonomic dysregulation in mutant rats

Despite the higher heart rates in HET rats (p<0.05; **Supplementary Figure 2A**), PR, QRS, J intervals, QT, and QTc remained unchanged indicating preserved ventricular conduction and repolarization (**Supplementary Figure 3**). These findings suggest that elevated heart rate arises primarily from enhanced sinoatrial node automaticity.

Twenty-four-hour telemetry trended elevated heart rates in HET and HOM compared to WT rats (**Figure 3A**). HRV analysis demonstrated marked autonomic imbalance: RMSSD and HF power were significantly reduced, LF power was increased, and the LF/HF ratio consequently elevated (**Figure 3B-E**). Poincaré plots showed a collapsed, cigar-shaped attractor in mutants (**Figure 3F**), consistent with loss of vagally mediated beat-to-beat variability and elevated basal sympathetic tone.^18^ Baroreflex sensitivity (BRS), an indicator of heart rate-blood pressure relationship, was significantly blunted (**Supplementary Figure 4**), In complementary experiments, direct recordings of sympathetic nerve activity during phenylephrine challenge further supported defective autonomic reflex regulation in mutant rats (Supplementary Figure 4).

**Figure 3.**
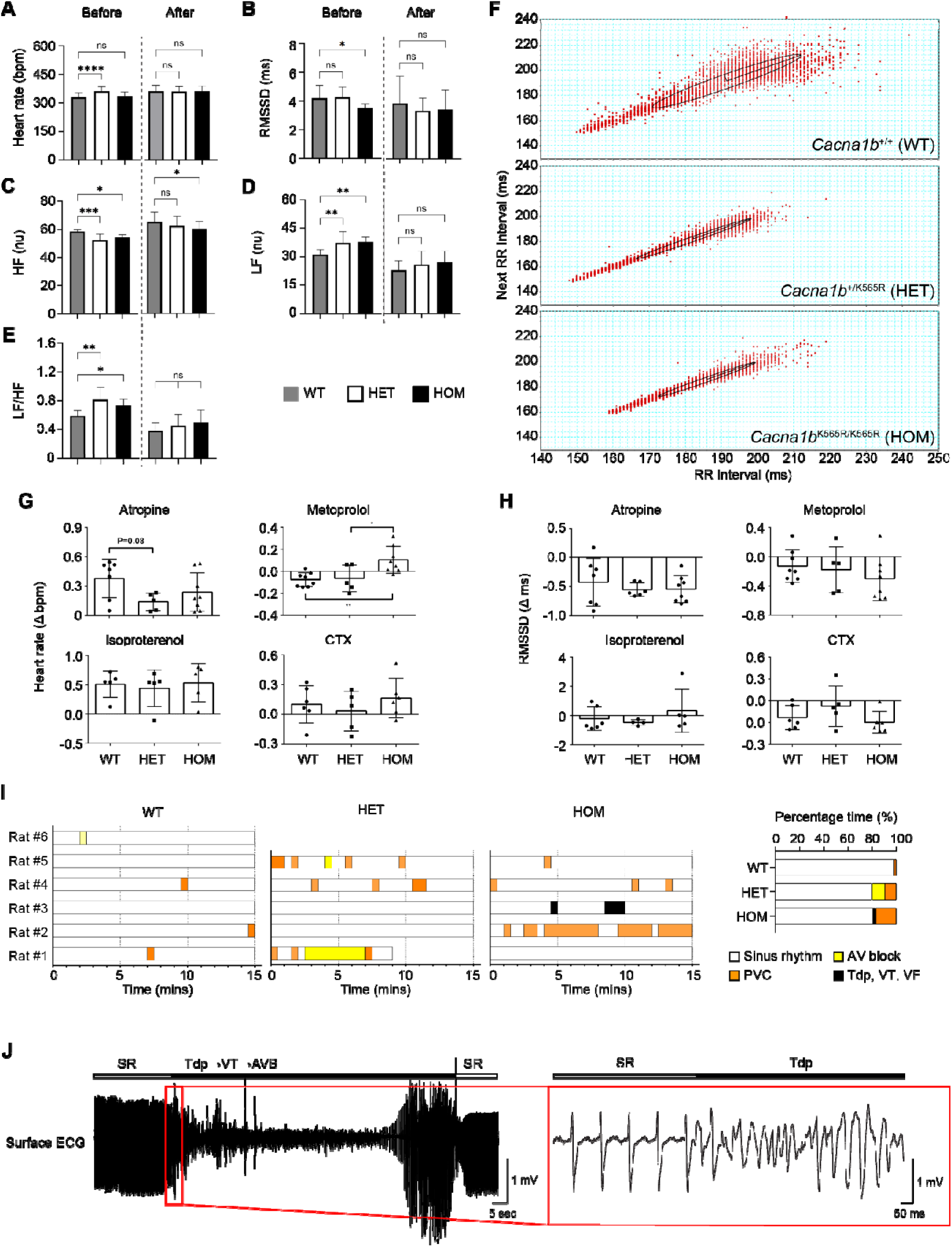
*Cacna1b* p.K565R knock-in rats exhibit constitutive sympathetic hyperactivity, autonomic dysregulation and arrhythmia risk *in vivo*. **(A-E)** Time- and frequency-domain heart rate variability (HRV) indices pre- and post-targeted pharmacological intervention (cilnidipine treatment, 10 mg/kg/day over 3 weeks). At baseline, mutant rats exhibit elevated heart rate **(A)**, diminished RMSSD **(B)**, reduced high-frequency (HF) **(C)**, elevated low-frequency (LF) power **(D)**, and resultant elevated LF/HF ratio **(E)**, suggestive of profound sympathetic dominance. This was effectively reversed and normalized by dual L/N-type calcium channel blockade across most indices (**A-E**). **(F)** Representative non-linear Poincaré plots of RR intervals derived from 24-hour ambulatory telemetry. HET and HOM mutants similarly display a tighter RR dispersion indicative of diminished beat-to-beat variability and vagal withdrawal. **(G-H)** Pharmacological dissection of the basal autonomic tone. Quantification of heart rate change (Λ1 bpm, **G**) and RMSSD change (Λ1 ms, **H**) in response to muscarinic blockade (atropine), β-adrenergic blockade (metoprolol), adrenergic provocation (isoproterenol), and specific N-type calcium channel blockade ( Ω -conotoxin GVIA, CTX). **(I)** Rhythm strip summaries from ambulatory ECG telemetry demonstrating a heightened susceptibility to isoproterenol-induced arrhythmogenesis in mutants. Observed arrhythmic events include premature ventricular contractions (PVCs), atrioventricular (AV) blocks, and life-threatening ventricular tachyarrhythmias characterized by Torsades de Pointes (TdP), ventricular tachycardia (VT), and ventricular fibrillation (VF). **(J)** Representative ECG trace with documented TdP ventricular arrhythmia in a HOM rat. Sample sizes of n=6/group (A-F), n=4-8/group (G and H), n=5-6/group (I). [Statistical analyses by one-way ANOVA with Tukey’s test (A-E, G-H).]

Pharmacologic interrogation further defined the altered autonomic baseline. Atropine produced a diminished heart rate rise (**Figure 3G**) and minimal RMSSD reduction (**Figure 3H**) in mutants, whereas metoprolol elicited an exaggerated bradycardic response. Isoproterenol induced hypersensitive tachycardia with marked HRV suppression (**Figures 3G-H**), but also triggered spontaneous arrhythmic events (**Figure 3I-J**). These included PACs/PVCs, atrioventricular block, and episodes of Torsades de Pointes-like polymorphic ventricular tachyarrhythmias.

Acute administration of the selective N-type calcium channel blocker Ω-conotoxin GVIA (CTX),^21^ rapidly normalized heart rates (Figure 3G) and restored vagal variability (Figure 3H) in mutant rats as in controls, confirming Ca_V_2.2 hyperactivity as the primary driver rather than compensatory remodeling. Chronic cilnidipine treatment (3 weeks), a dual N/L-type calcium channel blocker, effectively reversed the hyperadrenergic phenotype, normalizing heart rate and restoring LF, HF, and LF/HF indices to WT levels (**Figure 3A-E**). These findings demonstrate that Ca_V_2.2-mediated sympathetic hyperactivity is both a primary and reversible driver of autonomic pathology.

### Intracardiac electrophysiology: substrate for reentrant arrhythmias

Intracardiac electrophysiology revealed disruption of normal AV nodal decremental conduction in *Cacna1b* mutants. WT rats displayed smooth AV prolongation with decreasing S1-S2 coupling intervals. In contrast, HET and HOM rats demonstrated clear dual-pathway physiology, with abrupt AV “jumps” of ≥10 ms at critical coupling intervals. For example, in a representative HOM rat, shortening the S1–S2 coupling interval from 73 ms to 72 ms produced a paradoxical AV prolongation from 99 ms to 109 ms, indicating a fast-to-slow pathway switching (**Figure 4A**). Additionally, atrial echo beats were frequent (**Figure 4B**), and AVNRT was reproducibly inducible (**Figure 4C**), further confirming the presence of a functional re-entrant substrate. Dual pathway physiology occurred in 71.4% of HOM (5/7) and HET (5/7) rats, compared with 14.3% of WT (1/7). Echo beats were present in 100% of HOM (7/7) and 85.7% of HET (6/7), versus 28.6% of WT (2/7). Sustained AVNRT was inducible in 42.9% of HOM (3/7) and 14.2% of HET (1/7), but in none of the WT animals (0/7) (**Figure 4D**).

**Figure 4.**
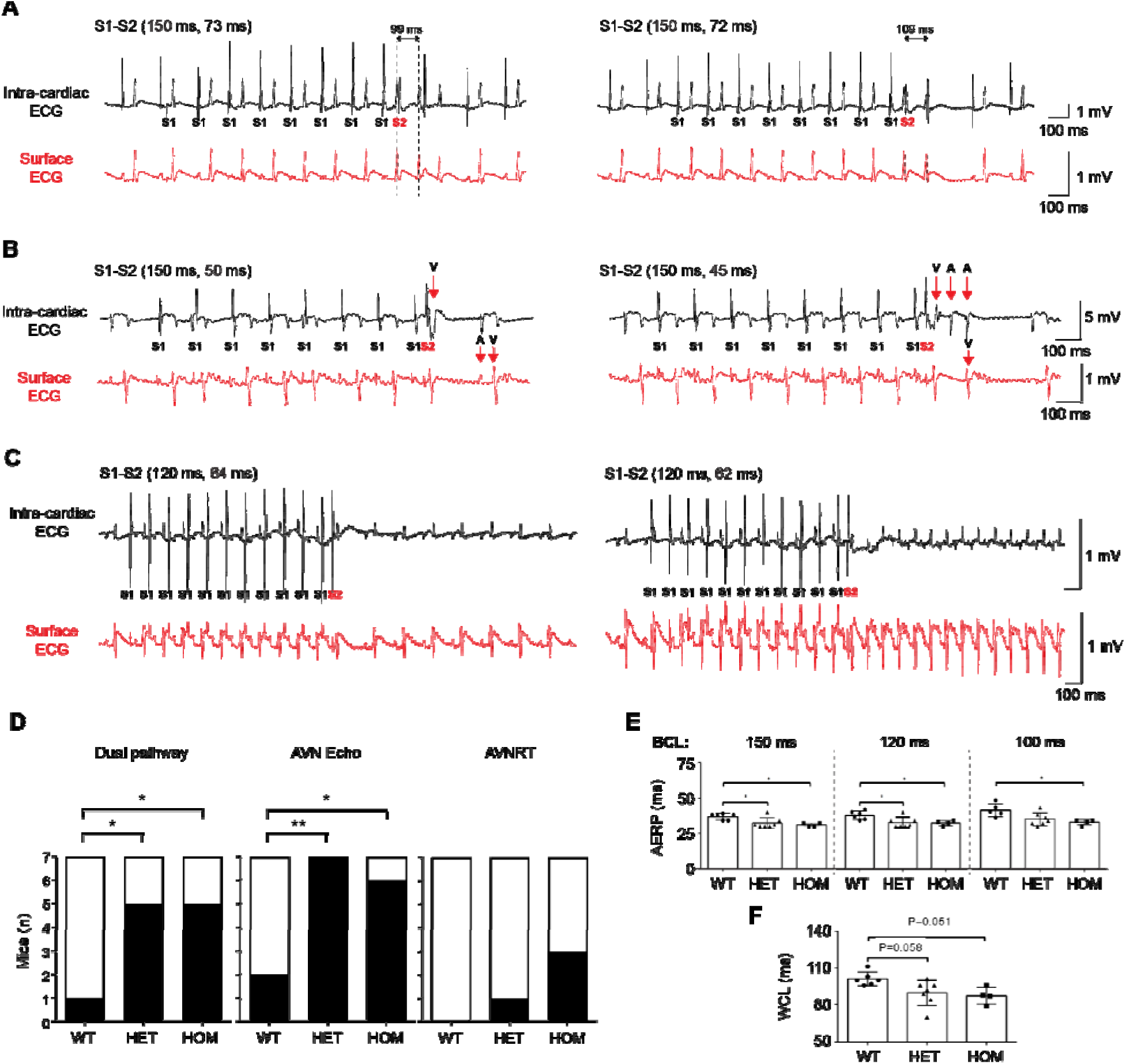
Intracardiac electrophysiological profiling reveals the classical triad of dual AV nodal pathways and AVNRT induction in mutant rats. **(A-C)** Representative intracardiac and corresponding surface ECGs during programmed electrical stimulation (PES) study in mutant rats. To demonstrate the conduction alterations, consecutive S1-S2 extrastimulus traces with decremented coupling intervals are displayed adjacent. These panels collectively illustrate the classical physiological manifestations of the dual AV nodal pathway. **(A)** AV nodal “jump” (a 1-ms decrement coupling interval triggers a disproportionate AH “jump” characterized by an interval prolongation 10 ms) confirming a shift from fast-to-slow AV nodal pathway conduction. **(B)** AV nodal echo beat (a critical premature atrial extrastimulus resulting in anterograde conduction via the slow pathway which retrogradely conducts up the fast pathway causing a single re-entrant echo beat (V-A-A-V sequence). **(C)** Induced sustained typical atrioventricular nodal re-entrant tachycardia (AVNRT) confirming a vulnerable arrhythmogenic substrate. **(D)** Number of individual subjects with reported incidence of the electrophysiological triad in mutant and WT rats. **(E)** Atrial effective refractory period (AERP) evaluated at basic cycle lengths (BCL) of 150, 120, and 100 ms, demonstrating significant shortening in mutant rats. **(F)** Wenckebach cycle length (WCL) measurements across genotypes. Biological replicates of n=6-7/group. Statistical analyses by Fisher’s exact test (D) and one-way ANOVA with Tukey’s test (E-F).

Consistent with the hyperadrenergic state (**Figure 3A-F**), AERP was significantly shortened in mutants (**Figure 4E**), whereas AVNERP and VERP were unchanged (**Supplementary Figure 5A-B**). Despite preserved nodal refractoriness, the WCL was significantly shorter in mutants (**Figure 4F**), indicating enhanced anterograde AV nodal conduction at rapid pacing rates. These findings demonstrate that the *Cacna1b* p.K565R mutation creates a robust reentrant substrate characterized by dual AV nodal pathways and shortened refractoriness, directly enabling reentrant tachycardia.^25,26^

### Heterologous overexpression in HEK293T cells unmasks intrinsic mutant Ca_V_2.2 gain-of-function

Whole-cell recordings in HEK293T cells expressing the minimal Ca_V_2.2 complex (α1B+β3) showed that p.K567R markedly increased inward Ba²⁺ currents across −30 to +60 mV (**Figure 5A-B**). Peak current density was significantly higher in mutant channels (**Figure 5C-E**), and currents were fully abolished by 1 μM ω conotoxin GVIA, confirming Ca_V_2.2 specificity. Voltage dependent activation and steady state inactivation were unchanged (**Figure 5F-G**), and minimal rundown ensured reliable pharmacological isolation (**Figure 5H**). These findings demonstrate that p.K567R produces a true intrinsic gain of function, likely by increasing functional channel surface density or open probability rather than altering gating kinetics.

**Figure 5.**
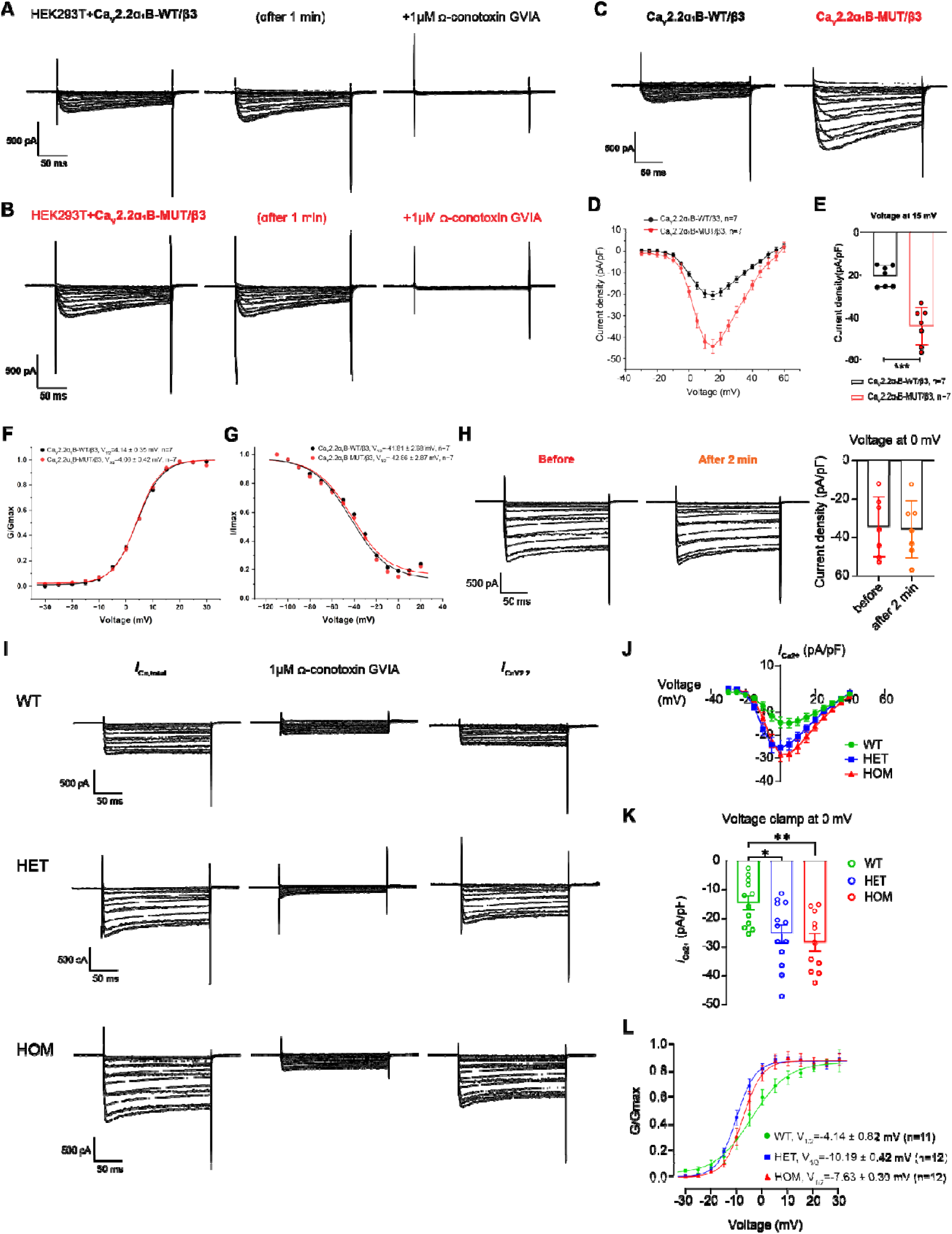
Biophysical characterization unveils an intrinsic GoF in mutant CaV2.2 channels and a calcium surge in native sympathetic neurons. **(A-G)** Heterologous expression of a recombinant CaV2.2 channel complex (pore-forming α1 and auxiliary β3 subunits) in HEK293T cells. **(A-C)** Representative whole-cell inward barium current (*I*_Ba2+_) traces in WT (**A**) and p.K567R (MUT, **B**) recorded at baseline at 1-minute intervals and after the application of 1 uM Ω-conotoxin GVIA (a specific N-type channel blocker) to identify Ca_V_2.2 currents and assess stability. **(C-E)** Representative whole-cell inward Ca_V_2.2 current traces in HEK293T co-expressing either WT and MUT CaV2.2 channel complex **(C)**, current-voltage (I-V) relationships across −30 to +60mV range in 5mV increments **(D)** and quantitation of peak current density elicited at 15 mV **(E)**, demonstrating an intrinsic GoF in the MUT channel. **(F-G)** Steady-state activation **(F)** and inactivation **(G)** curves reveal preserved voltage-gating kinetics. **(H-L)** Whole-cell patch-clamp recordings from native superior cervical ganglion (SCG) neurons isolated from rats to validate impact on the autonomic environment. **(H)** Representative total Ca^2+^ current traces in SCG neurons of 1-to-3-week-old Sprague-Dawley rats in response to holding potential was -90 mV and depolarizing test voltages ranging from −30 to +40 mV in 5 mV increments before and after a 2-minute interval patch clamp without -conotoxin GVIA application demonstrating stable calcium current amplitude (n=7). **(I)** Representative total calcium current (*I*_Ca,total_) before and after application of 1 uM Ω -conotoxin GVIA. The subtraction of the two derives N-type calcium current (*I*_CaV2.2_) traces across WT, HET, and HOM genotypes. **(J-K)** Current-voltage (I-V) relationship **(J)** and peak Ca_V_2.2 current density (at 0 mV) reveals a surge in mutant sympathetic neurons. **(L)** Comparison of voltage-dependent activation kinetics in mutant SCG neurons indicates a hyperpolarizing shift towards enhanced voltage sensitivity. Data expressed as mean ± SE Biological replicates of n=7/group (A-H), n=10-12/group (I-L). Statistics (student t-test (D, E, H), one-way ANOVA with Tukey’s multiple comparisons (J, K), G/Gmax and I/Imax curves were analyzed by fitting to a Boltzmann function (F, G, L).

### Knock-in rats’ SCG neurons show constitutive Ca_V_2.2 hyperactivity and increased surface density

To validate physiological relevance, we recorded N-type calcium currents (*I*_Ca,N_) from SCG neurons of *Cacna1b* p.K565R knock-in rats. The SCG is the major sympathetic ganglion innervating the cardiac conduction system.^44^ Mutant neurons displayed a robust, gene-dosage-dependent increase in current amplitude across the physiological voltage range (**Figure 5I-K**). Peak current density increased by ∼84% in HOM mutants (−29.60±3.34 vs. −16.06±2.29 pA/pF; p<0.01) and was intermediate in HET (−26.51±3.08 pA/pF; p<0.05), consistent with autosomal dominant inheritance (**Figure 5J-K**).

Gating analysis showed a slight depolarizing shift in V_1/2_ in HET and HOM neurons (HET: - 10.19±0.42; HOM: −7.63±0.30 vs. WT: −4.14±0.82 mV; **Figure 5L**), a shift that would theoretically reduce activation. The paradoxical increase in macroscopic current despite this shift strongly supports increased functional channel surface density as the dominant mechanism. Thus, p.K565R drives constitutive sympathetic hyperactivity by increasing the number of active Ca_V_2.2 channels at the membrane.

### Single-cell transcriptomics reveals a hyper-glutamatergic sympathetic state

Despite the pronounced electrophysiological gain of function, mRNA and protein levels of *CACNA1B* and other Ca² channel genes (*CACNA1A, CACNA1E, CACNA1C, CACNA1G*) were unchanged across SCG (**Supplementary Figure 6A-D**), stellate ganglion, adrenal gland, and atrioventricular node (data not shown).

Unsupervised UMAP clustering of SCG tissue scRNA-seq (45,761 cells) identified 10 major populations, including glutamatergic (Glut) and noradrenergic (NA) neurons (**Figure 6A-B**). Mutant SCG displayed a near doubling of Glut neurons (WT: 122; HET: 215). Within this subset, we observed marked gene upregulation of presynaptic exocytotic machinery, Synaptotagmin-1 (*Syt1*; primary Ca^2+^ sensor for exocytosis) and Synaptosomal-Associated Protein 25 (*Snap25*; core SNARE component)^45^ (**Figure 6C**). Enrichment analyses of Glut_HET_UP genes showed strong activation of synaptic signaling, chemical synaptic transmission, synaptic vesicle cycle, and regulation of synaptic organization (**Figure 6D**). Canonical autonomic markers (ChAT, TH, NET) remained unchanged (**Supplementary Figure 6A**), indicating preserved noradrenergic identity despite enhanced excitatory drive.

**Figure 6.**
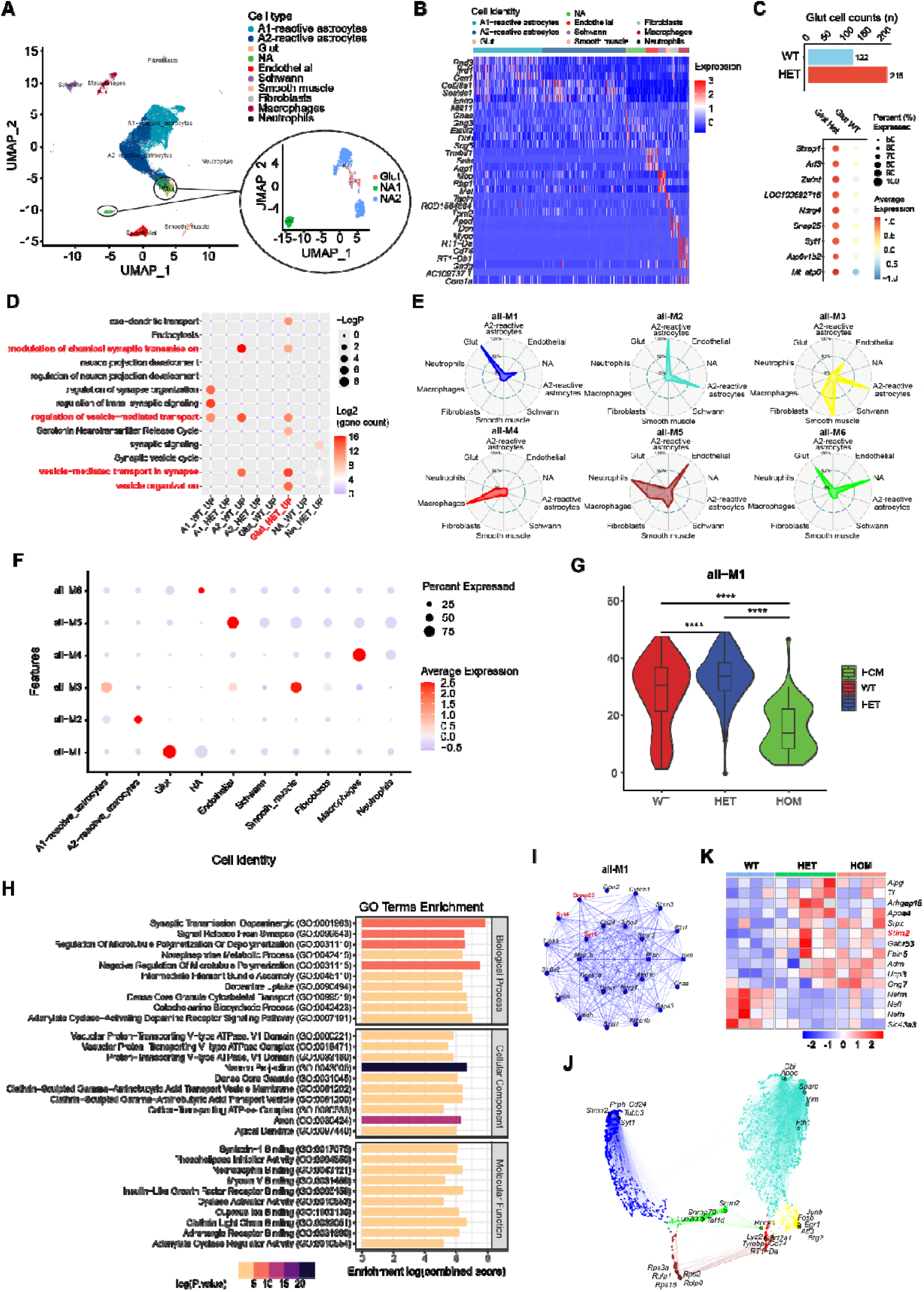
Single-cell transcriptomics reveal a mutant-specific hyper-glutamatergic neuronal state of the SCG. **(A)** Uniform Manifold Approximation and Projection (UMAP) of cells from WT and mutant SCG. Magnified sub-clustering of the neuronal compartment identifies distinct glutamatergic (Glut) and noradrenergic (NA1, NA2) subpopulations. **(B)** Heatmap displaying the top cellular marker genes across differential cell types within the ganglion. **(C)** Cell count and dot plot illustrating the expansion of the Glut neuronal population in HET animals with increased expression of presynaptic components such as *Syt1* and *Snap25* (key genes in synaptic vesicle cycling and fusion machinery). **(D)** Gene Ontology (GO) enrichment analysis (bubble plot) highlights robust upregulation of biological processes of “Synaptic Neurotransmitter Release” and “Synaptic Organization” in mutant neurons. **(E-J)** High-dimensional WGCNA pinpoints a *Syt1/Snap25*-driven synaptic hyperactivity module governing the neuro-cardiac axis. **(E)** Radar charts mapping the eigengene expression distribution across six distinct gene modules (M1-M6), with M1 exhibiting specificity and robust enrichment for Glut cells. **(F)** Dot plot summarizing the fractional expression and average expression levels of the identified modules across SCG cell lineages. **(G)** Violin plot demonstrating allele-dose-dependent escalation in the M1 module eigengene score in HET and HOM mutant rats compared to WT controls. **(H)** Gene Ontology (GO) enrichment analysis of the M1 module, underscoring “Synaptic Vesicle Exocytosis” and “Signal Release from Synapse” as the fundamental pathological drivers. **(I)** Protein-protein interaction (PPI) network topology of the M1 module, definitively pinpointing *Syt1*-*Snap25* as core intra-modular hubs orchestrating the hyperadrenergic state. **(J)** UMAP plot visualizing spatial distribution of M1 activity across the ganglion. **(K)** TMT-based proteomic heatmap of differentially expressed proteins from WT, HET, and HOM SCG tissue. Mutants demonstrate increased neurosecretory and synaptic proteins (*Stim2*, *Adm*, *Gng7*, and *Gabrb3*) together with reduced structural and metabolic proteins (*Nefl*, *Nefm*, *Nefh*, *Arhgap15*, and *Ucp3*), supporting coordinated remodeling of SCG function. Biological replicates of n=1-2/group (A-J) and n=5/group (K).

High dimensional WGCNA identified a mutation associated module (M1) with elevated eigengene expression in mutant Glut neurons (**Figure 6E-G**). M1 was enriched for neurotransmitter synthesis, vesicular exocytosis, catecholamine metabolism, and synaptic release pathways. Cellular component analysis revealed enrichment in dense core granules, V ATPase complexes, and neuronal projections (**Figure 6H**). Network topology identified *Syt1* and *Snap25* as central hubs (**Figure 6I–J**), linking increased CaV2.2 influx to enhanced vesicle fusion.

TMT-proteomics corroborated these findings: mutant SCG exhibited upregulation of the calcium sensor STIM2, the neuroendocrine peptide adrenomedullin (ADM), and synaptic modulators (G Protein Subunit Gamma 7, Gng7; Gamma-Aminobutyric Acid Type A Receptor Subunit Beta3, Gabrb3), alongside downregulation of the structural neurofilament triplet (light, NEFL; medium, NEFM; heavy, NEFH), the cytoskeletal regulator Rho GTPase Activating Protein 15 (ARHGAP15), and mitochondrial uncoupling protein-3 (UCP3) (**Figure 6K**). Together, these data reveal a coordinated molecular program that enhances neurosecretory output and remodels cytoskeletal and metabolic architecture.

### Neuro-cardiac crosstalk: metabolic reprogramming of the AV node

UMAP clustering of AV node scRNA-seq (88,748 cells) resolved 12 populations, including central sinoatrial node (cSAN) pacemaker cells, transitional (Tz) cells, epicardial neurons, and structural lineages (**Figure 7A-B**). Sub-clustering identified four cardiomyocyte subtypes, including a distinct Tz population bridging cSAN and working myocardium (**Figure 7C-E**).

**Figure 7.**
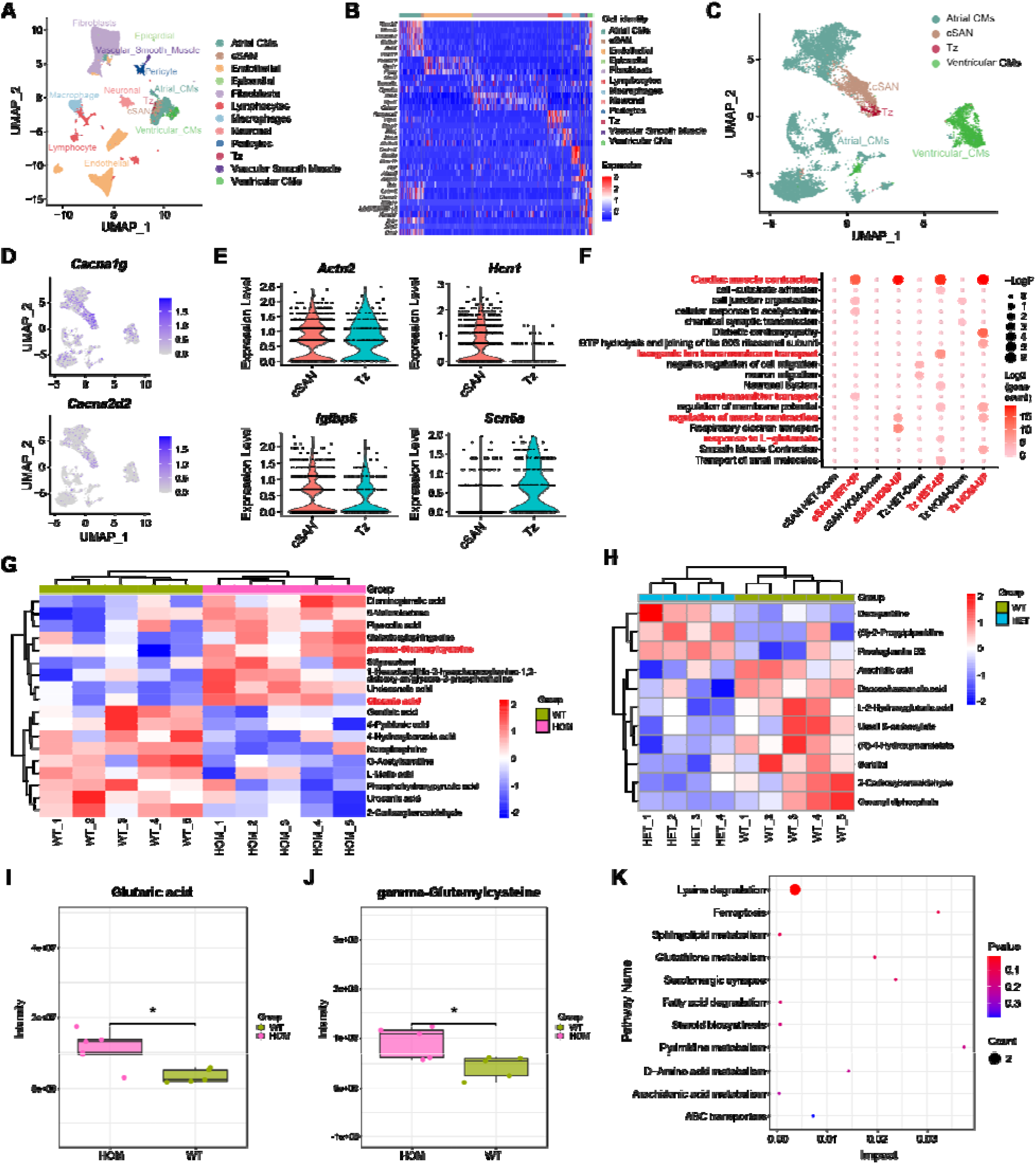
Single-cell transcriptomics of the atrioventricular node reveal a mutant-specific glutamatergic response. **(A)** UMAP plot of isolated cells from the atrioventricular node (AVN) tissue. The clustering resolves the cardiac and non-myocyte lineages, delineating specialized conduction system subpopulations including transitional cells (Tz) and compact Sinoatrial/Atrioventricular node cells (cSAN). **(B)** Heatmap illustrates canonical lineage-specific marker gene profiles reflecting 12 differential AVN cell populations. **(C)** UMAP sub-clustering of the broader cardiomyocyte population further delineates four distinct myocardial and conduction system subtypes. **(D)** UMAP feature plots projecting localized expression of critical pacemaker channel genes, *Cacna1g* and *Cacna2d2*, predominantly within the cSAN and associated conduction clusters. **(E)** Violin plots depicting the expression of *Actn2*, *Hcn1*, *Igfbp5*, and *Scn5a* in pacemaker and transitional cells. **(F)** Gene Set Enrichment Analysis (GSEA, bubble plot) showing enrichment of the “response to L-glutamate” pathway in mutant Tz cells, consistent with an increased glutamatergic sympathetic input from the SCG supporting a neuro-cardiac association between sympathetic activation and downstream AV nodal remodeling. **(G–K)** Non-targeted metabolomic profiling of AVN tissue from WT, HET, and HOM rats. **(G)** OPLS-DA and hierarchical clustering heatmap analyses demonstrating distinct global metabolomic profiles across genotypes. **(H)** Heatmap of representative differential metabolites showing mutation-associated remodeling of the AVN metabolome. **(I–J)** Box plots showing elevated γ-glutamylcysteine **(I)** and glutaric acid **(J)** levels in mutant AVN tissue. **(K)** KEGG enrichment pathway analysis identifies glutathione metabolism, fatty acid degradation, and lysine degradation as major altered pathways in HOM animals. Collectively, these findings suggest local metabolic reprogramming and oxidative stress in the mutant AVN. Biological replicates of n=2/group (A-F) and n = 5/group (G-K).

Mutant Tz cells (Tz_HET_UP and Tz_HOM_UP) exhibited marked enrichment of neural-associated programs, including response to L-glutamate, chemical synaptic transmission, neuronal system, and cardiac muscle contraction (Figure 7F), indicating that transitional nodal cells are active downstream targets of excessive sympathetic–glutamatergic input from the SCG. Mutant Tz and cSAN populations also upregulated pathways regulating membrane potential and ion transmembrane transport, consistent with increased electrophysiological vulnerability within the nodal microenvironment. Concurrent enrichment of respiratory electron transport pathways further suggested compensatory mitochondrial hyperactivation under chronic neurogenic stress. Together with the metabolomic evidence of glutathione-pathway activation, these findings support a coupled electrophysiological and metabolic remodeling program in transitional nodal cells that may increase susceptibility to reentrant conduction.

Untargeted metabolomic analyses demonstrated clear separation of WT, HET, and HOM AVN samples, indicating broad reprogramming of the nodal metabolome (Figure 7G–H). Among the altered metabolites, mutant AVN tissue showed accumulation of γ-glutamylcysteine, the rate-limiting precursor for glutathione synthesis, together with increased glutaric acid levels (**Figure 7I–J**), mapping to glutathione metabolism.^46^ KEGG pathway enrichment of upregulated metabolites in the homozygous group further prioritized glutathione metabolism, fatty acid degradation, and lysine degradation (Figure 7K). These findings are consistent with local metabolic remodeling in the mutant AV node and suggest the presence of a metabolically stressed microenvironment.

### Molecular and structural remodeling of the sympathetic synapse

To link enhanced Ca² entry to neurotransmission, we examined presynaptic proteins in SCG. Both HET and HOM ganglia showed elevated SYT1 and SNAP25 protein levels (**Figure 8A-B**). This protein-level increase was further validated at the cellular level. Immunofluorescence confirmed increased SYT1 and SNAP25 intensity in cultured SCG neurons (**Figure 8C**), indicating assembly of a “hyper competent excitosome” that enhances vesicle fusion.^45,47^ TEM of SCG synapses showed preserved architecture and vesicle density (**Supplementary Figure 9A–B**), indicating that hyperactivity arises from molecular, not structural, changes.

**Figure 8.**
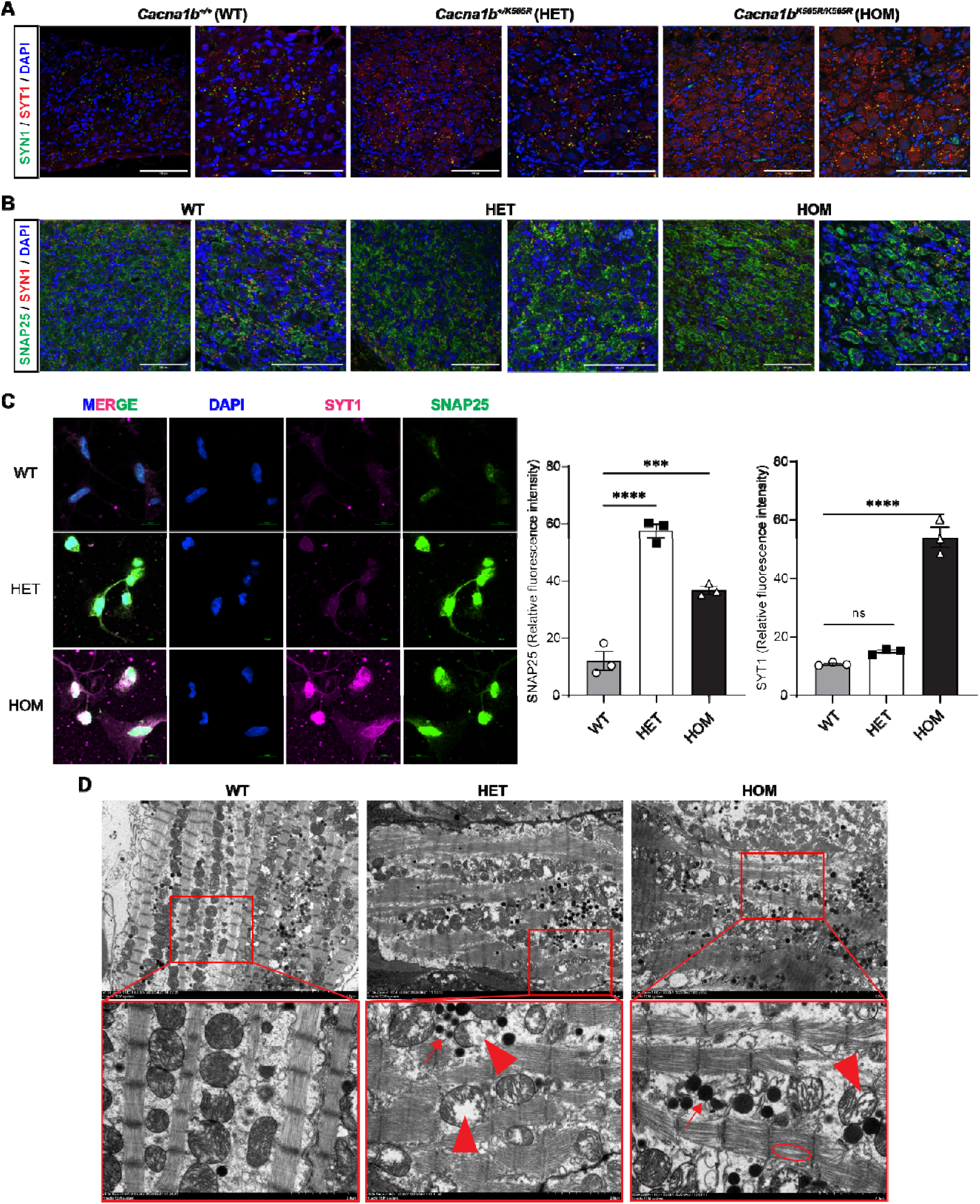
Molecular upregulation of the presynaptic fusion machinery drives target-specific ultrastructural degradation along the neuro-cardiac axis. **(A-B)** Representative immunofluorescence images in SCG tissues from WT, HET, and HOM rats for expression of SYT1 **(A)** and SNAP25 **(B)** colocalized to SYN1 expression as part of the core presynaptic fusion machinery. **(C)** Representative immunofluorescence confocal microscopy of cultured primary SCG neurons displaying SYT1 (magenta) and SNAP25 (green) colocalization. Mutant neurons display elevated fluorescence intensity of SNAP25 in both mutants but upregulated SYT1 in HOM only. Pooled n=4 biological replicates per group per experiment (conducted thrice separately). Statistical analyses by one-way ANOVA. **(D-E)** Transmission electron microscopy (TEM) revealed marked morphological divergence between the upstream ganglionic source and the downstream cardiac target. **(D)** Mutant SCG neurons maintain a structurally preserved synaptic architecture and stable vesicle density, reflecting a primary *functional*—rather than structural—overdrive. **(E)** Conversely, downstream AVN cardiomyocytes exhibit profound subcellular remodeling and structural degradation driven by relentless sympathetic bombardment. Pathological hallmarks of this localized metabolic stress include extensive mitochondrial damage (characterized by pathological enlargement, reduced matrix electron density, disrupted cristae, and focal vacuolization), abundant accumulation of glycogen granules and lipid droplets (lipotoxicity), alongside early manifestations of myofibrillar dissolution (myolysis). Biological replicates of n=3 rats/group (D, E).

In contrast, AV nodal tissue exhibited enlarged mitochondria with reduced matrix density, disrupted cristae, focal vacuolization, glycogen accumulation, lipid droplets, and mild myofibrillar dissolution (**Figure 8D**). These features indicate profound metabolic stress consistent with chronic sympathetic overdrive.

## Discussion

### A neurogenic mechanism underlying familial AVNRT

Using zebrafish overexpression and a *Cacna1b* knock in rat, we provide the first definitive evidence that *CACNA1B* (CaV2.2) is a disease gene for neurogenic familial AVNRT. Unlike classical cardiac channelopathies that directly alter cardiomyocyte electrophysiology, the p.K567R variant drives arrhythmogenesis via sympathetic hyperactivation, reshaping the neuro cardiac interface to generate a functional dual-pathway substrate for reentry.

A key advance of this study is the demonstration that a presynaptic Ca² channel gain-of-function mutation drives sympathetic overdrive and secondary AV nodal remodelling. Single cell and multi omics data converge on a common axis: the mutation enhances presynaptic excitability by expanding a glutamatergic sympathetic subpopulation and upregulating vesicle fusion machinery (SYT1/SNAP25), thereby amplifying neurotransmitter release and imposing sustained sympathetic pressure on the AV node. Downstream, transitional (Tz) cells exhibit metabolic stress, mitochondrial activation, and altered ion handling pathways,^48^ collectively forming a vulnerable reentrant substrate. The normalization of elevated heart rate and impaired heart rate variability following cilnidipine treatment indicates that the autonomic disturbance is functionally maintained and pharmacologically reversible. Although repeat intracardiac electrophysiological testing was not performed after treatment, these findings nonetheless support the sympathetic neuro-cardiac junction as a potentially tractable therapeutic target.^8,49^

### Ca_V_2.2 GoF as the molecular driver

*CACNA1B* encodes the α1 subunit of the presynaptic N-type voltage-gated Ca^2+^ channel (Ca_V_2.2), enriched in sympathetic neurons, where it governs Ca^2+^-dependent neurotransmitter release.^14^ Prior *Cacna1b* knockout studies linked loss-of-function to sympathetic hypofunction and impaired baroreflex sensitivity,^50^ but no GoF phenotype has been described.

In this present study, Zebrafish overexpression of the novel *CACNA1B* p.K567R variant revealed a robust chronotropic effect in the developing embryo, and the orthologous knock-in rat exhibited constitutive sympathetic overdrive—elevated heart rate, impaired HRV, and blunted vagal reflexes—effectively reversed by cilnidipine, confirming that N-type channel hyperactivity is causal.^20^ Electrophysiology demonstrated dual-pathway physiology, AV nodal echo beats, and inducible AVNRT—rare in rodents—indicating that Ca_V_2.2 GoF is sufficient to generate an AV nodal reentrant substrate. Although repeat intracardiac electrophysiological testing was not performed after cilnidipine treatment, the normalization of elevated heart rate and impaired heart rate variability indicates that this autonomic phenotype is functionally maintained and pharmacologically reversible. Together, these findings identify CaV2.2 GoF as an upstream molecular driver linking presynaptic calcium-channel dysfunction to sustained sympathetic overactivity and downstream AV nodal arrhythmogenic remodeling.

### The Ca_V_2.2–SYT1–glutamate axis driving sympathetic hyperactivity

To further define the molecular basis of this sympathetic hyperactivity, we next examined the downstream presynaptic machinery linking CaV2.2 activity to neurotransmitter release. This is mechanistically important because SYT1, the canonical fast Ca² sensor, is a low affinity, high threshold sensor requiring steep Ca² microdomains to trigger exocytosis.^51^ Enhanced Ca² influx through mutant Ca_V_2.2 likely saturates SYT1 C2 domains, accelerating vesicle fusion and expanding the readily releasable pool.^52,53^

snRNA-seq identified *Slc17a6*^+^ (VGLUT2) glutamatergic sympathetic neurons as the epicenter of the hyperactive gene module (M1). Sympathetic neurons co release glutamate and norepinephrine to facilitate rapid excitatory transmission,^54^ and our data indicate that p.K567R selectively amplifies this glutamatergic arm. Sn-RNA sequencing further showed that Tz cells upregulate neural associated signaling (“response to L glutamate”, “chemical synaptic transmission”)^55,56^ and mitochondrial pathways, indicating direct sensing of sympathetic and glutamatergic input. AV nodal metabolomics revealed accumulation of γ-glutamylcysteine and glutaric acid, with enrichment of glutathione metabolism and fatty acid degradation, consistent with clinical associations between elevated glutamate and sympathetic overactivity.^41^ Ultrastructural mitochondrial abnormalities and mild myofibrillar dissolution further support a metabolically stressed, oxidatively injured microenvironment. Beyond telemetry-based HRV abnormalities, blunted baroreflex responses provide independent physiological evidence of sympathetic dysregulation in the mutant model.

Collectively, these findings support a model of neurogenic excitotoxicity in which Ca_V_2.2 GoF drives excessive glutamate and catecholamine release, remodeling AV nodal tissue, shortening refractoriness, and establishing a reentry-amenable substrate.

### Clinical implications: targeting the neuro-cardiac axis

Identifying *CACNA1B* as the first synaptic gene linked to familial AVNRT reframes the disorder as autonomic dependent reentry rather than a purely anatomical phenomenon. This suggests that the genetic architecture of idiopathic PSVT extends beyond cardiomyocytes ion channels to genes regulating autonomic signaling, synaptic transmission, and vesicle cycling. Clinically, such patients may exhibit catecholamine sensitive arrhythmias that respond poorly to vagal maneuvers but favorably to sympathetic modulation.

Therapeutically, these data provide proof of concept for precision targeting of the neuro cardiac axis. Catheter ablation eliminates the circuit but not the upstream driver; β blockers blunt receptor signaling but do not reduce neurotransmitter release. In contrast, cilnidipine directly suppresses N type Ca² dependent exocytosis, normalizes autonomic tone, and abolishes the functional reentrant substrate. For patients with autonomic genetic drivers, N type calcium channel blockade offers a rational, mechanism based, non invasive therapeutic strategy.

### Limitations

Several limitations merit consideration. First, although the *Cacna1b* knock in rat recapitulates key features of human AVNRT, species specific differences in autonomic regulation and AV nodal microanatomy may limit direct translation. Second, while multi omics localized the neurogenic romodeling to the AV node, its microscopic size precluded direct patch clamp recordings from transitional (Tz) cells; the electrophysiological consequence of localized oxidative stress therefore requires future validation, ideally using spatial transcriptomics or high resolution optical electrophysiology to map sympathetic–Tz synaptic interactions. Third, although the SCG emerged as the dominant sympathetic driver, the contribution of the intrinsic cardiac nervous system (ganglionated plexi) remains undefined. Finally, while cilnidipine abolished arrhythmia inducibility, its dual N-/L type blocking profile complicates in vivo separation of presynaptic N type inhibition from postsynaptic L type effects on nodal conduction.

### Conclusion

We identify a *CACNA1B* (Ca_V_2.2) GoF mutation as a novel genetic driver of familial AVNRT. By enhancing presynaptic N type calcium influx, p.K567R hyperactivates glutamatergic sympathetic neurons; this sustained autonomic overactivation induces metabolic and oxidative stress within atrioventricular nodal Tz cells, creating a vulnerable substrate for reentrant conduction. Targeted N-/L type calcium channel blockade with cilnidipine effectively neutralizes this neuro cardiac axis, restoring autonomic balance and eliminating the functional substrate for AVNRT. These findings establish a neurogenic paradigm for AVNRT, shifting the disorder from a static, anatomically determined entity to a dynamic, autonomically driven arrhythmia, and open the door to precision medicine for autonomic related and neurocardiogenic arrhythmias.

## Author Contributions

X.L. conceptualized the study, supervised the entire project, and analyzed the clinical pedigree data. R.L., X.C., and C.Z. contributed equally to this work. R.L. generated the *Cacna1b* knock-in rat model, characterized the arrhythmia phenotype, and drafted the original manuscript. X.C. performed in vivo electrophysiological studies, programmed electrical stimulation (PES), and cellular patch-clamp recordings. C.Z. conducted WES, scRNAseq, and bioinformatic analyses of the multi-omics data. X.W. established the zebrafish model and performed morphological and heart rate assessments. Q.S. acquired and analyzed echocardiographic data. S.L. performed optical mapping experiments and data processing. H.D. conducted molecular biology assays (Western blotting, qPCR) and immunofluorescence staining. L.Y. contributed to experimental design and data interpretation. W.L. assisted with data interpretation, figure preparation, critically revised the manuscript, and provided linguistic editing. All authors read and approved the final manuscript.

---

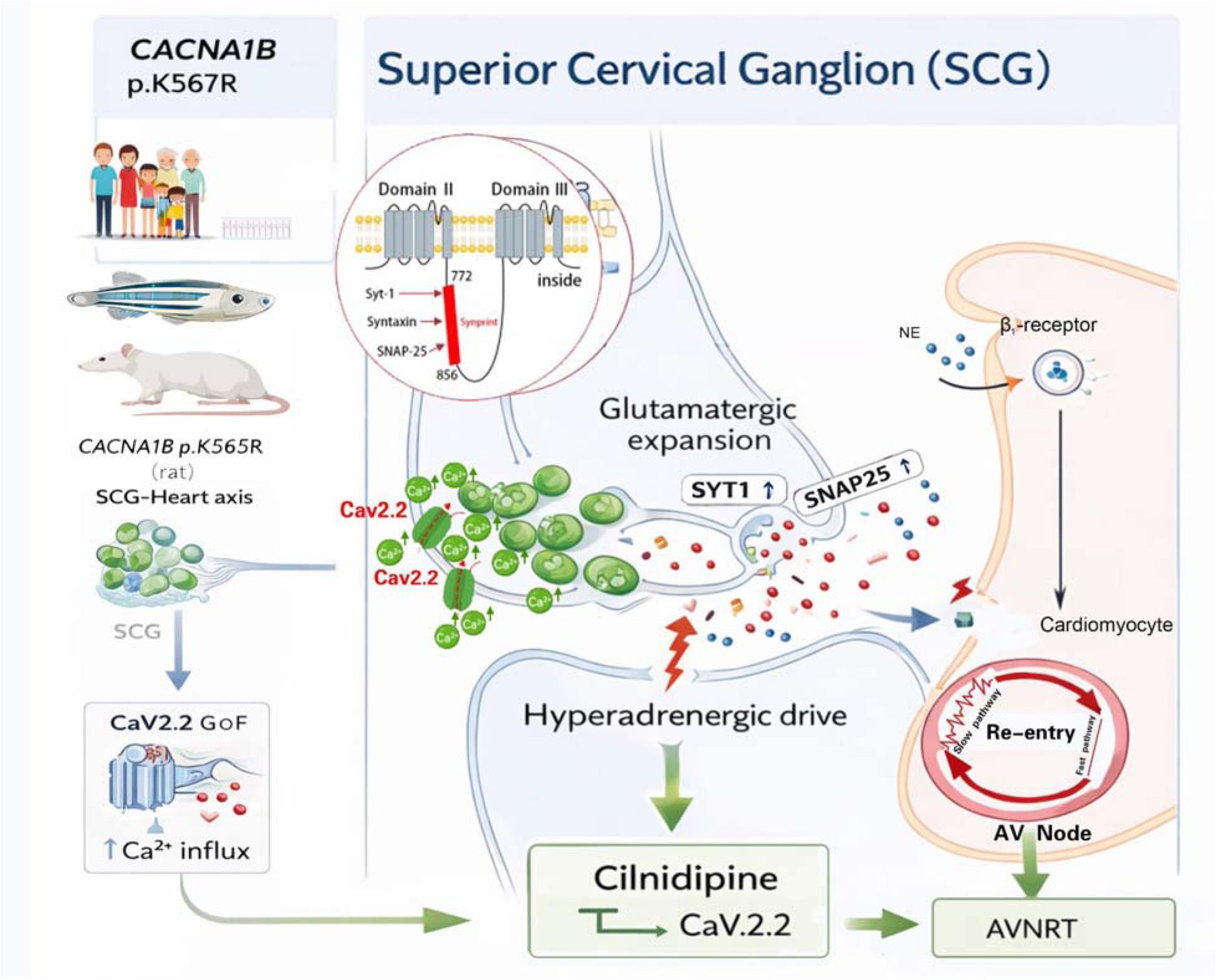

A GoF mutation (p.K567R) in the *CACNA1B* (Cav2.2) channel orchestrates a profound hyperadrenergic state within the superior cervical ganglion (SCG). By amplifying N-type calcium influx, this variant specifically upregulates the presynaptic fusion machinery (SYT1/SNAP25) within the glutamatergic neuronal compartment, unleashing a relentless neurochemical bombardment upon the heart. This chronic sympathetic overdrive drives extensive metabolic and subcellular remodeling of the atrioventricular node (AVN), thereby establishing the discontinuous electrophysiological substrate prerequisite for AVNRT. Crucially, targeted pharmacological blockade with cilnidipine restores autonomic homeostasis and abrogates this arrhythmogenic neuro-cardiac axis.

## Supplementary Tables

**Supplementary Table S1.**
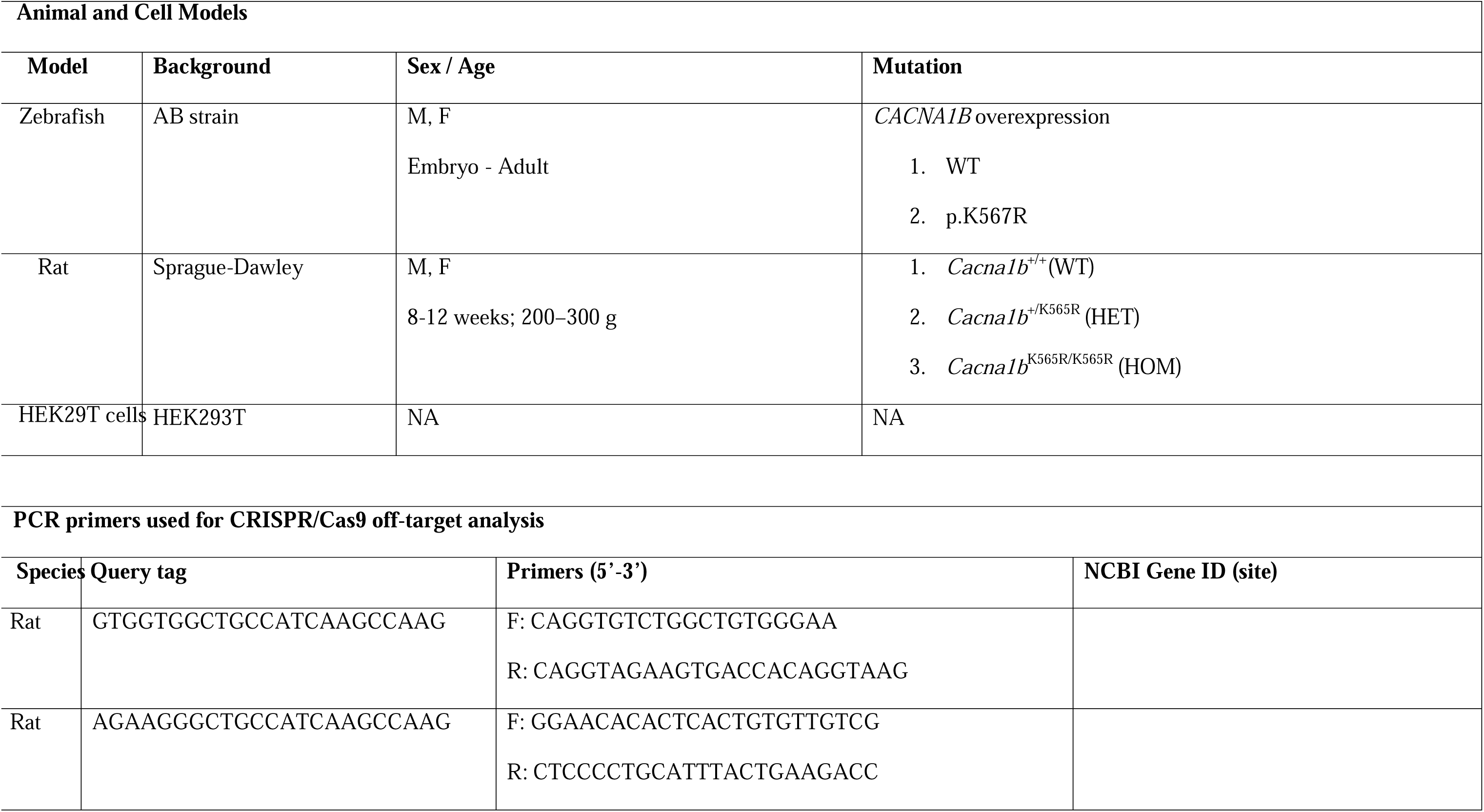
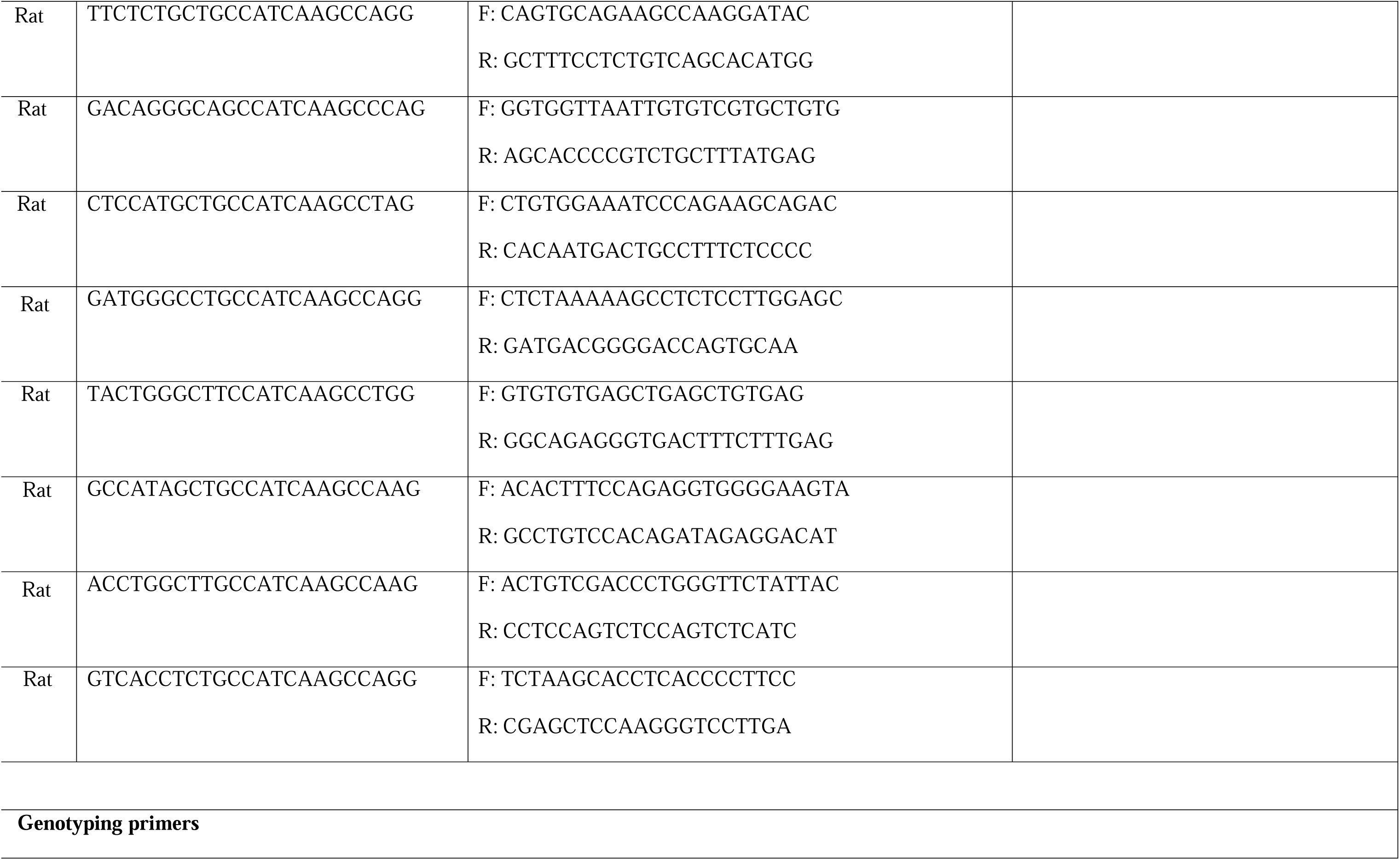
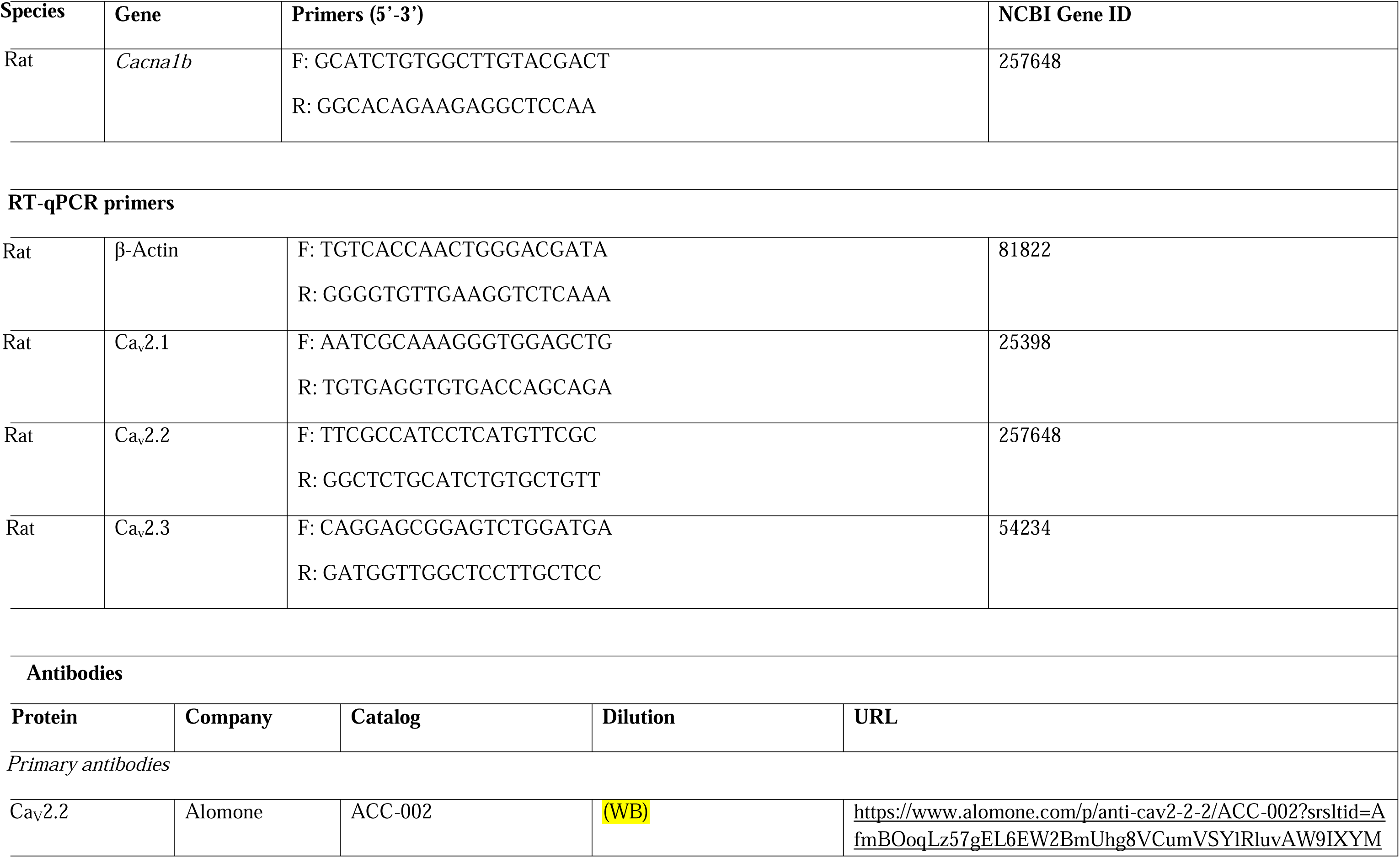
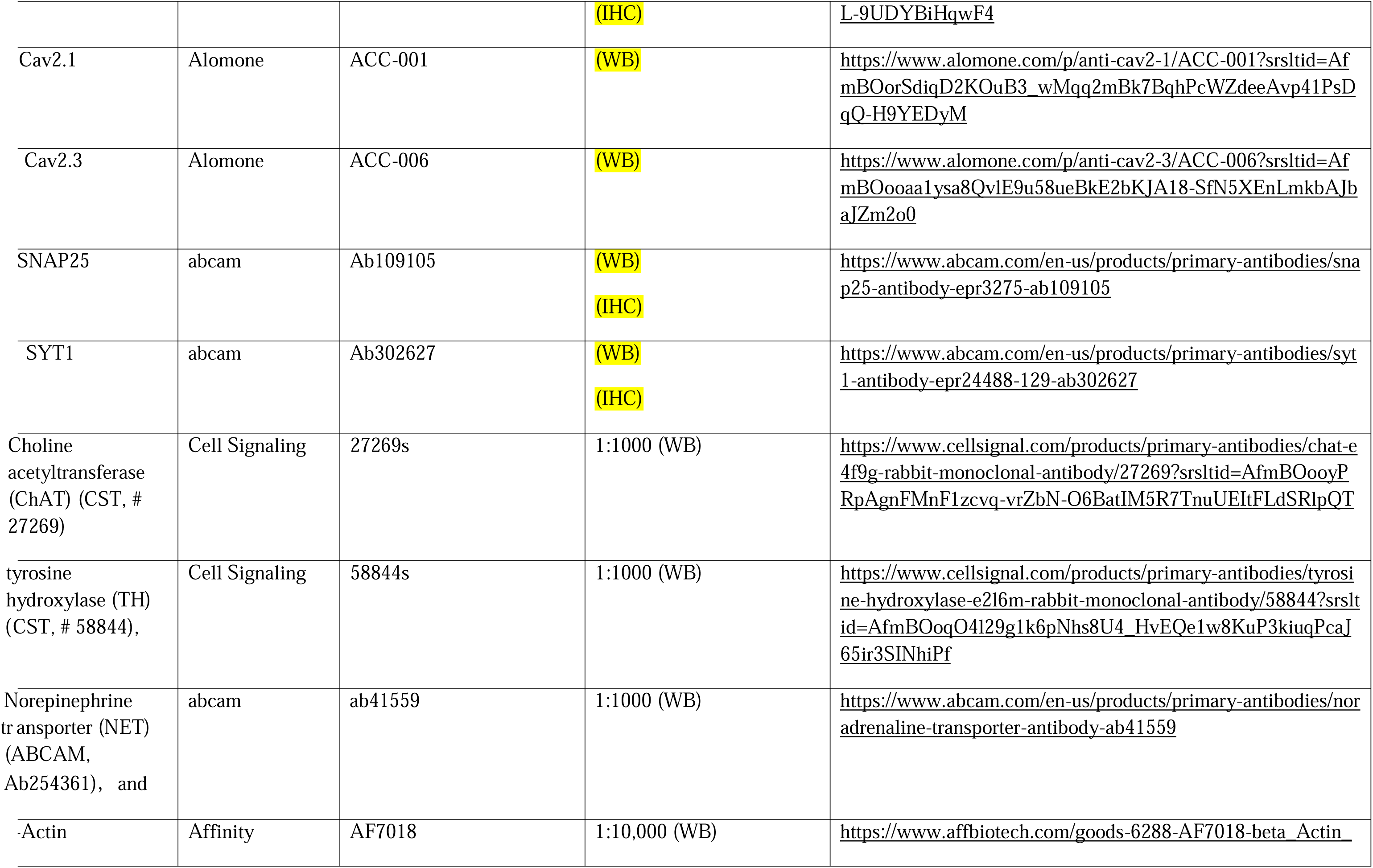
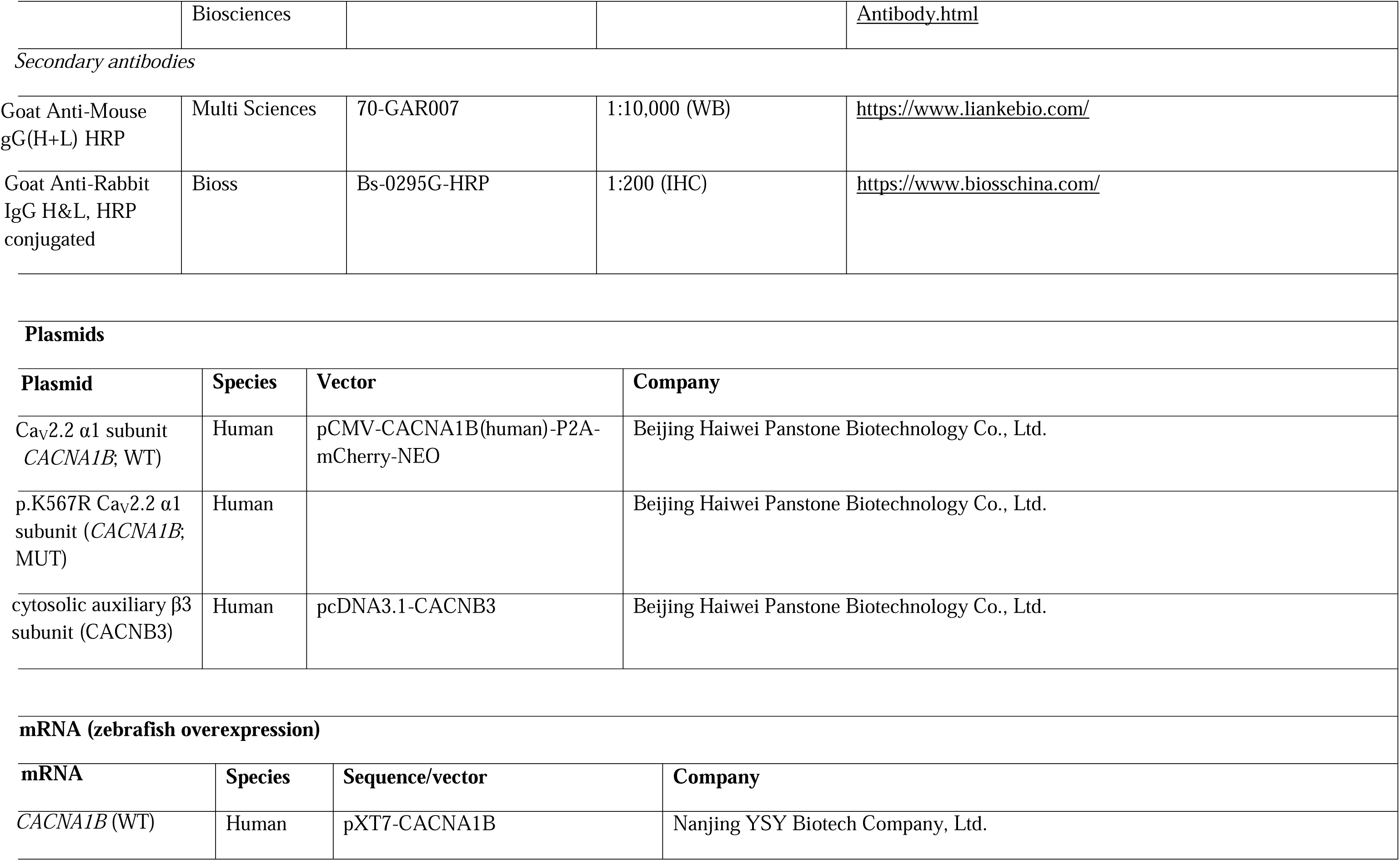
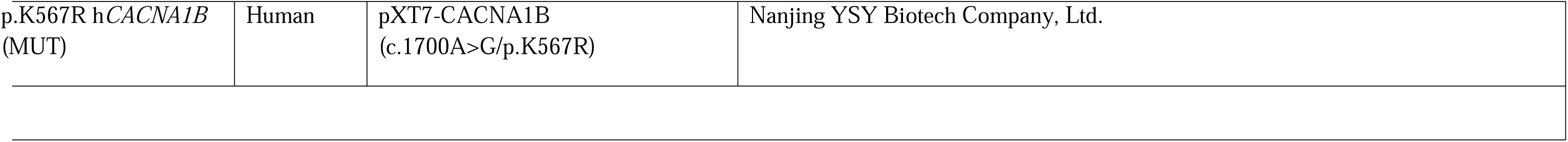
Major Resources Table.

**Supplementary Table S2.** Clinical features and ACMG classification of *CACNA1B* variants identified in 86 sporadic AVNRT patients.

| Sample ID | cDNA Change | Protein Change | Location (Exon) | Functional Domain | MAF (Global) | MAF (East Asian) | In Silico Prediction | Conservation | ACMG Criteria | ACMG Classification |
| --- | --- | --- | --- | --- | --- | --- | --- | --- | --- | --- |
| J754 | c.493G>A | p.Gly165Ser | Exon 3 | Ion Transport Domain | 4.14E-05 | 0 | Possible pathogenic | Yes | PM2, PP3 | Uncertain Significance (VUS) |
| AR249, AR619 | c.6744G>C | p.Gln2248His | Exon 47 | C-terminus | 0.0006 | 0.0066 | Possible pathogenic | Yes | PM2, PP3 | Uncertain Significance (VUS) |

**Notes:** Variants were classified according to the American College of Medical Genetics and Genomics (ACMG) guidelines. MAF, Minor Allele Frequency; VUS, Variant of Uncertain Significance. PM2, pathogenic moderate criterion 2 (absent or extremely rare in population databases); PP3, pathogenic supporting criterion 3 (multiple *in silico* computational algorithms support a deleterious effect on the gene/protein).

**Supplementary Table S3.**
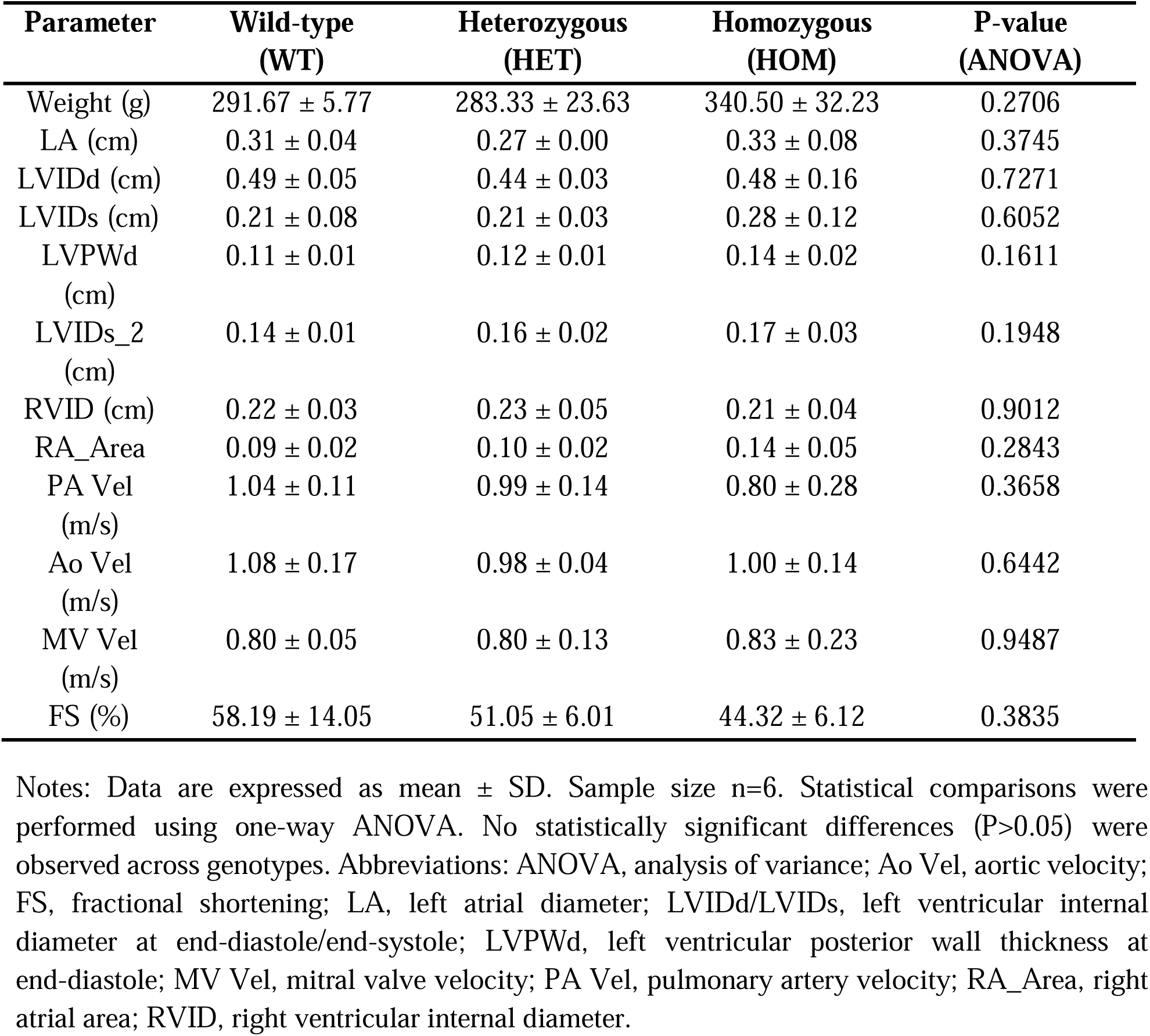
Baseline echocardiographic parameters across different *Cacna1b* genotypes in rats.

| Parameter | Wild-type<br>(WT) | Heterozygous<br>(HET) | Homozygous<br>(HOM) | P-value<br>(ANOVA) |
| --- | --- | --- | --- | --- |
| Weight (g) | 291.67 ± 5.77 | 283.33 ± 23.63 | 340.50 ± 32.23 | 0.2706 |
| LA (cm) | 0.31 ± 0.04 | 0.27 ± 0.00 | 0.33 ± 0.08 | 0.3745 |
| LVIDd (cm) | 0.49 ± 0.05 | 0.44 ± 0.03 | 0.48 ± 0.16 | 0.7271 |
| LVIDs (cm) | 0.21 ± 0.08 | 0.21 ± 0.03 | 0.28 ± 0.12 | 0.6052 |
| LVPWd<br>(cm) | 0.11 ± 0.01 | 0.12 ± 0.01 | 0.14 ± 0.02 | 0.1611 |
| LVIDs_2<br>(cm) | 0.14 ± 0.01 | 0.16 ± 0.02 | 0.17 ± 0.03 | 0.1948 |
| RVID (cm) | 0.22 ± 0.03 | 0.23 ± 0.05 | 0.21 ± 0.04 | 0.9012 |
| RA_Area | 0.09 ± 0.02 | 0.10 ± 0.02 | 0.14 ± 0.05 | 0.2843 |
| PA Vel<br>(m/s) | 1.04 ± 0.11 | 0.99 ± 0.14 | 0.80 ± 0.28 | 0.3658 |
| Ao Vel<br>(m/s) | 1.08 ± 0.17 | 0.98 ± 0.04 | 1.00 ± 0.14 | 0.6442 |
| MV Vel<br>(m/s) | 0.80 ± 0.05 | 0.80 ± 0.13 | 0.83 ± 0.23 | 0.9487 |
| FS (%) | 58.19 ± 14.05 | 51.05 ± 6.01 | 44.32 ± 6.12 | 0.3835 |

Notes: Data are expressed as mean ± SD. Sample size n=6. Statistical comparisons were performed using one-way ANOVA. No statistically significant differences (P>0.05) were observed across genotypes. Abbreviations: ANOVA, analysis of variance; Ao Vel, aortic velocity; FS, fractional shortening; LA, left atrial diameter; LVIDd/LVIDs, left ventricular internal diameter at end-diastole/end-systole; LVPWd, left ventricular posterior wall thickness at end-diastole; MV Vel, mitral valve velocity; PA Vel, pulmonary artery velocity; RA\_Area, right atrial area; RVID, right ventricular internal diameter.

## Supplementary Figures

**Supplementary Figure 1.**
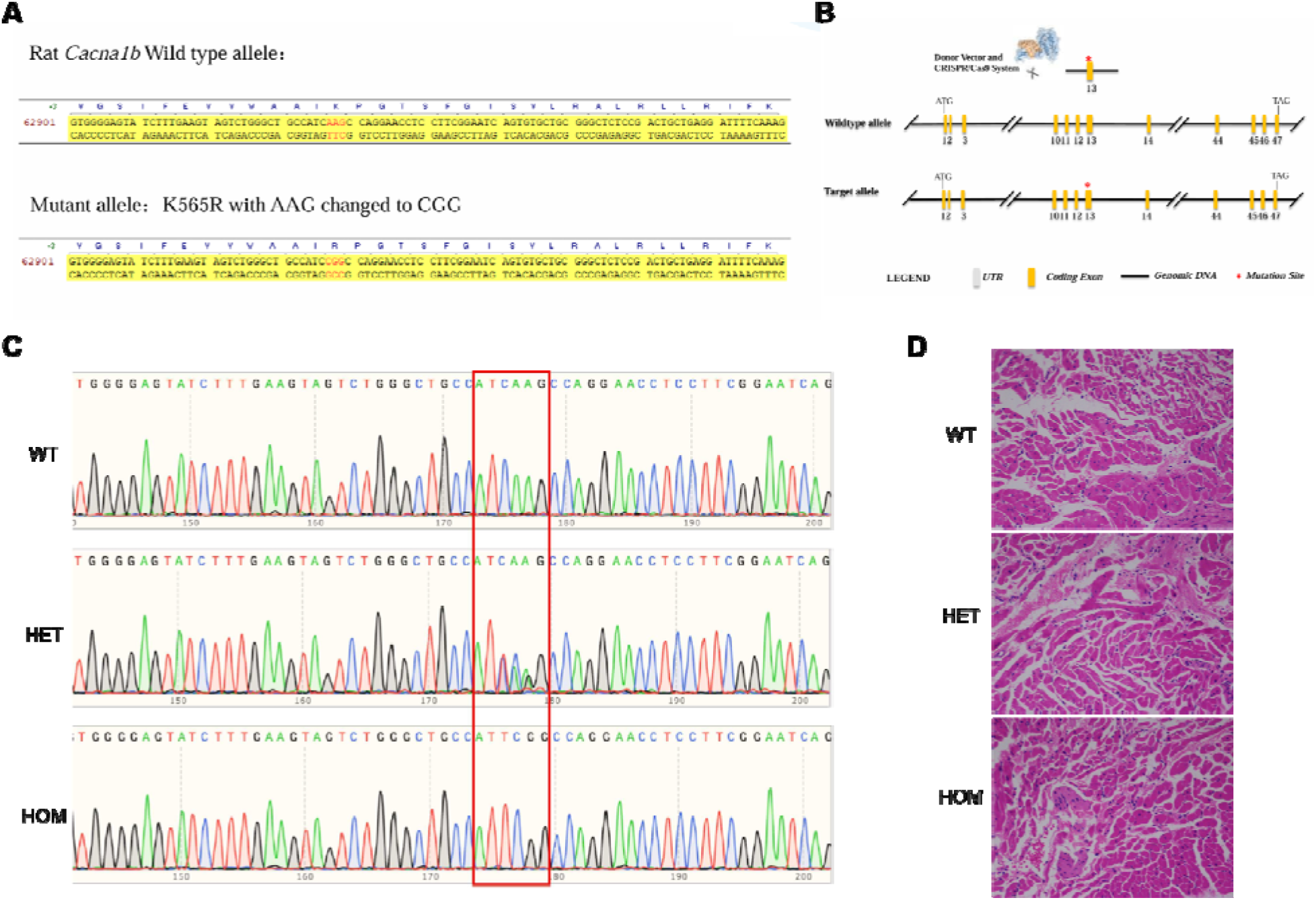
Generation and molecular validation of the *Cacna1b* p.K565R knock-in rat model. **(A)** Schematic representation of the CRISPR/Cas9-mediated genome editing strategy. A single-stranded oligodeoxynucleotide (ssODN) donor was utilized to precisely introduce the pathogenic point mutation (AAG-CGG) into exon 13 of the endogenous rat *Cacna1b* locus. **(B)** Representative PCR-based genotyping results from genomic DNA, effectively differentiating WT, HET, and HOM mutant rats. **(C)** Representative Sanger sequencing chromatograms validating the precise nucleotide substitution at the targeted genomic locus, definitively confirming the genetic fidelity of each generated genotype. **(D)** Representative hematoxylin and eosin (H&E) stained cardiac cross-sections from WT, HET, and HOM rats. The images demonstrate well-preserved baseline cardiac cytoarchitecture, confirming the absence of overt macroscopic or microscopic structural remodeling prior to arrhythmogenesis.

**Supplementary Figure 2.**
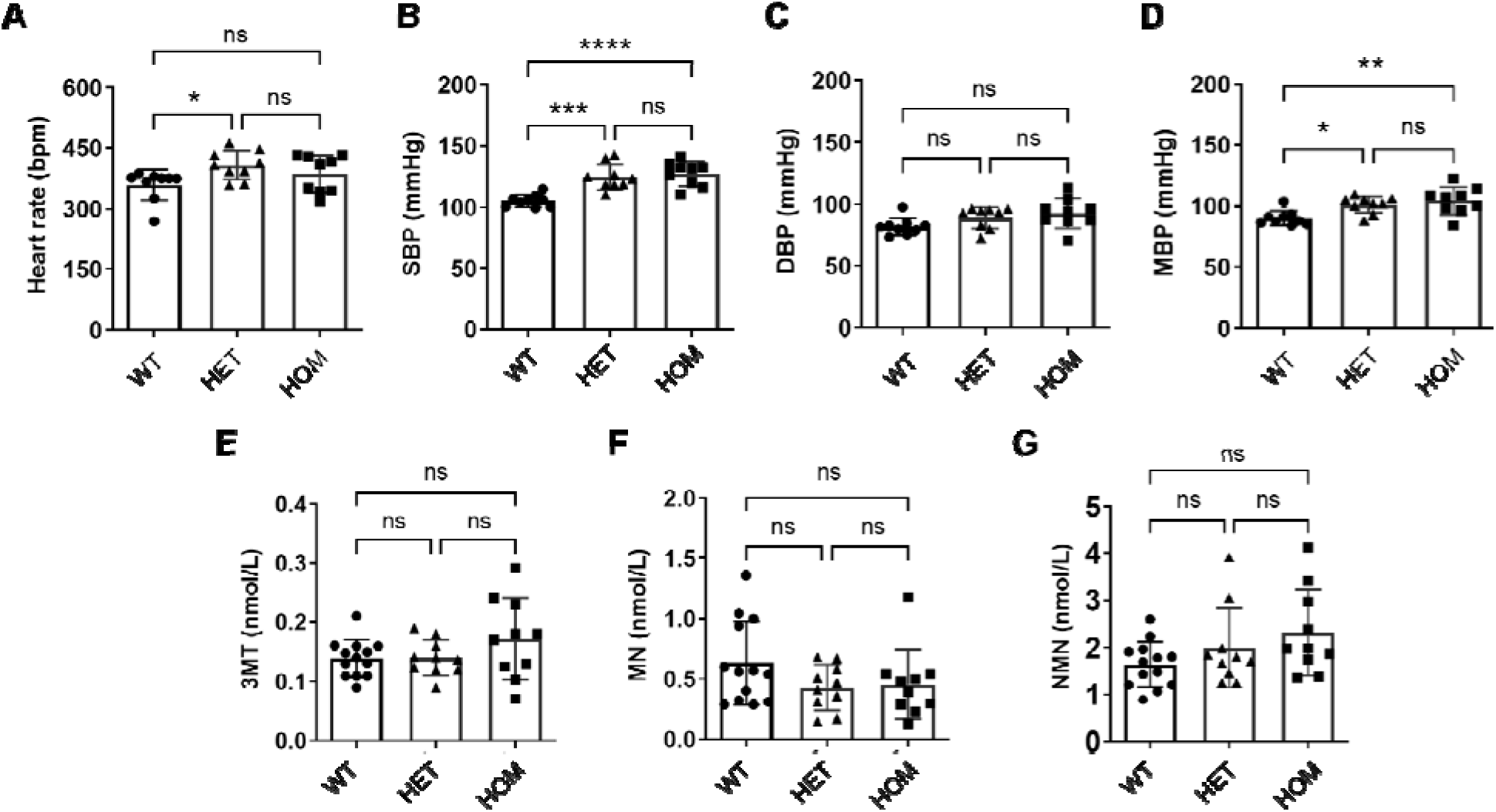
Hemodynamic profiles and circulating catecholamine levels in the *Cacna1b* p.K565R knock-in rat model. **(A-D)** *In vivo* telemetric assessment of heart rate (**A**), systolic blood pressure (SBP, **B**), diastolic blood pressure (DBP, **C**), and mean arterial pressure (MAP, **D**) in conscious, freely moving *Cacna1b* WT, HET, and HOM rats. **(E-G)** Quantitative ELISA profiling of stable serum catecholamine metabolites to evaluate systemic sympathetic tone. The panels detail the circulating concentrations of 3-methoxytyramine (3-MT, a dopamine metabolite; **E**), metanephrine (MN, an epinephrine metabolite; **F**), and normetanephrine (NMN, a norepinephrine metabolite; **G**). Sample size of n=9/group (A-D), n= 10-13/group (D-F). Data are presented as mean ± SD. Statistical significance was determined using one-way ANOVA (*P<0.05, **P<0.01, ***P< 0.001, ****P<0.0001).

**Supplementary Figure 3.**
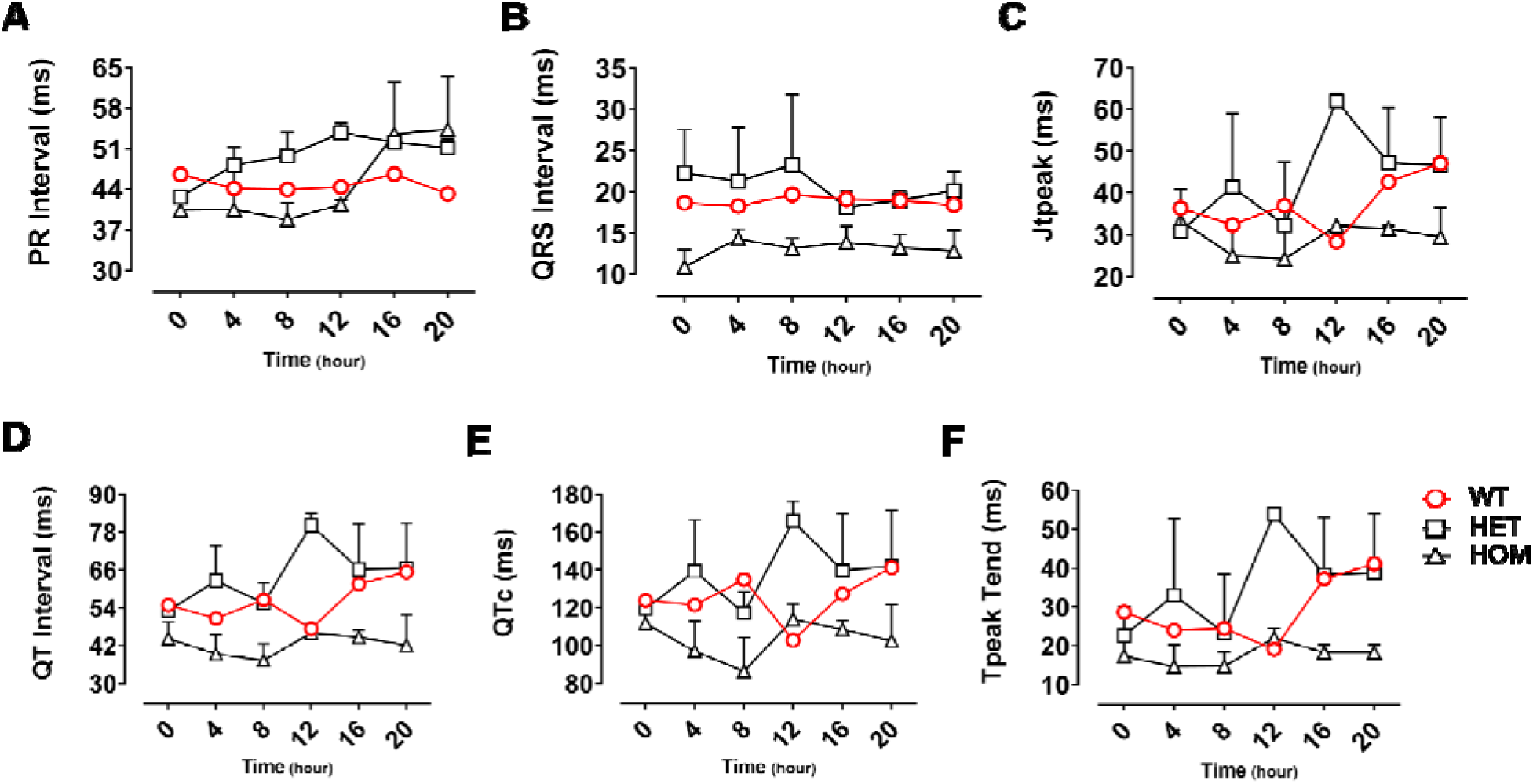
Comprehensive 24-hour ambulatory surface electrocardiographic (ECG) profiling across *Cacna1b* genotypes. **(A-F)** Quantitative assessment of standard surface ECG indices over a continuous 24-hour monitoring period in conscious *Cacna1b* WT, HET, and HOM rats. Evaluated parameters include PR interval **(A)**, QRS duration **(B)**, Jt peak interval **(C)**, uncorrected QT interval **(D)**, rate-corrected QT (QTc) **(E)**, and the Tpeak-Tend interval **(F)**. In alignment with the established hyperadrenergic state, mutant rats display persistent resting tachycardia. Indices of ventricular depolarization (QRS duration) and global repolarization (QTc and Tpeak-Tend) remain unaffected. This specific preservation of ventricular electrophysiology firmly substantiates that the proarrhythmic substrate is localized and neurogenically driven, rather than reflecting a generalized myocardial channelopathy. Data are presented as mean ± SEM. Biological replicates of n=??/group. Statistical analyses by one-way ANOVA with Tukey’s test.

**Supplementary Figure 4.**
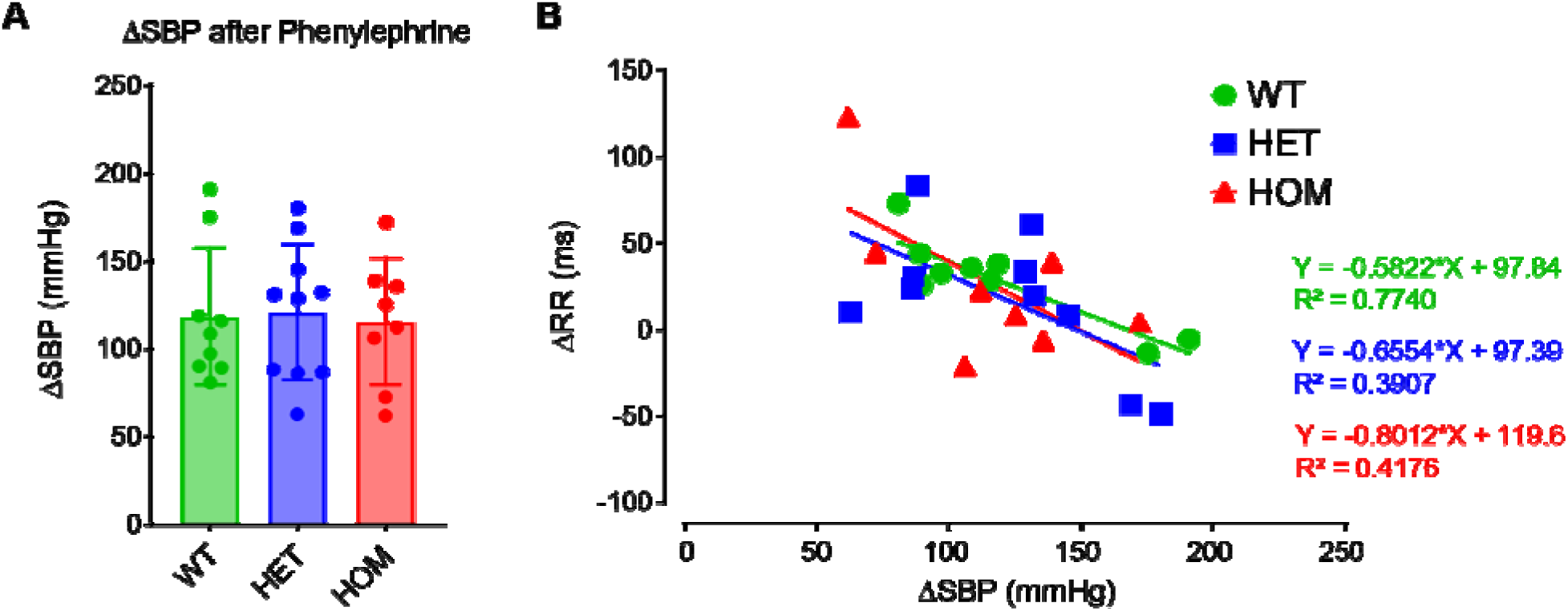
Blunted baroreflex sensitivity (BRS) reveals profound autonomic dysregulation in *Cacna1b* knock-in rats. (A) Assessment of baroreflex responses across WT, HET and HOM *Cacna1b* genotypes with the quantitative analysis of changes in systolic blood pressure (SBP) following a pharmacological challenge with phenylephrine (200 µg/kg iv). (B) Linear regression analysis mapping the beat-to-beat relationship between systolic blood pressure and the subsequent RR interval to derive spontaneous baroreceptor sensitivity (BRS) response. Both HET and HOM mutants exhibit a significantly flattened regression slope (P=0.0533 and P=0.0834 respectively) compared to WT controls which demonstrate a significant non-zero slope (P=0.0018). This attenuation in mutants suggests a blunted vagal reflex arc driven by the mutation. Biological replicates of n=8-10/group. Statistical analyses were determined by one-way ANOVA with Tukey’s test **(A)** and simple linear regression **(B)**.

**Supplementary Figure 5.**
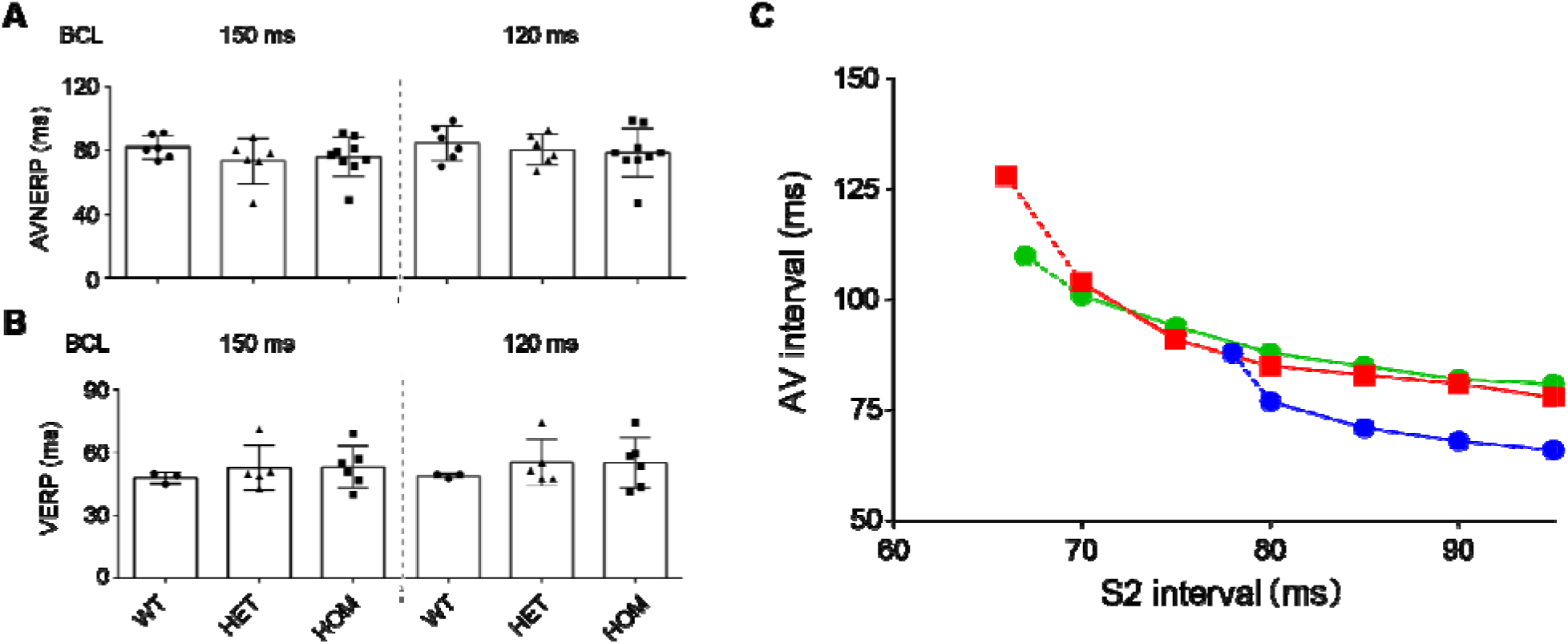
Invasive intracardiac electrophysiology reveals preserved basal atrioventricular nodal and ventricular refractoriness across *Cacna1b* genotypes. **(A-B)** Quantitative assessment of the atrioventricular nodal effective refractory period (AVNERP; **A**) and ventricular effective refractory period (VERP; **B**) utilizing invasive intracardiac programmed electrical stimulation. Measurements were acquired at resting basic cycle lengths (BCL) of 150 ms and 120 ms in *Cacna1b* WT, HET, and HOM mutants. Due to inconsistent 1:1 capture of AVNERP and VERP at BCL of 100 ms, we excluded further analyses at the faster pacing rates. Notably, no statistically significant differences in fundamental refractoriness were observed among the groups under these basal pacing conditions. This crucial preservation of intrinsic electrophysiological properties indicates that the arrhythmogenic substrate is driven by dynamic sympathetic overdrive rather than constitutive myocardial ion channel remodeling. **(C)** Composite AV nodal conduction curves plotting the AH interval against the S1-S2 coupling interval. Data are expressed as mean ± SEM. Biological replicates of n=3-9/group. Statistical analyses by one-way ANOVA with Tukey’s test.

**Supplementary Figure 6.**
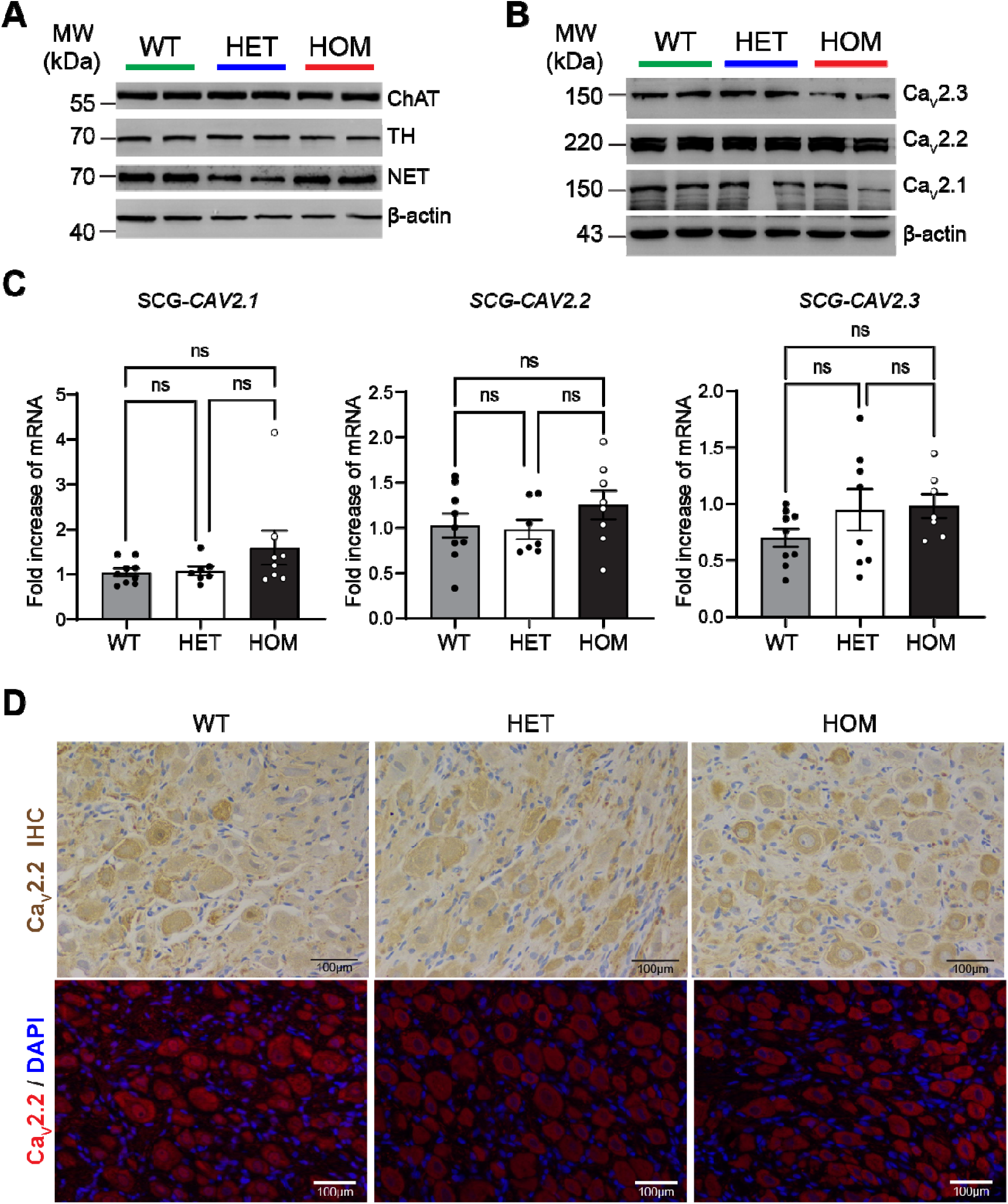
Stable protein expression and preserved subcellular distribution of Ca_V_2.2 in the superior cervical ganglion (SCG) across *Cacna1b* genotypes. **(A-B)** Quantitative protein expression profiling of native superior cervical ganglion (SCG) tissues. **(A)** Representative immunoblots evaluating total Ca_V_2.2 protein levels in SCG homogenates from WT, HET, and HOM rats. Canonical autonomic lineage markers, including choline acetyltransferase (ChA; cholinergic) and tyrosine hydroxylase (TH; noradrenergic), and noradrenaline transporter (NET) were co-assessed to ensure ganglionic phenotypic stability, with β-actin serving as the loading control. **(B)** Corresponding densitometric quantification reveals that the total steady-state expression of the Ca_V_2.2 channel remains strictly conserved, showing no significant differences across genotypes. **(C)** Representative immunohistochemical (IHC) staining of SCG tissue sections. These data suggests preserved spatial subcellular localization and overall abundance of Ca_V_2.2 in mutant rats. Data expressed as mean ± SEM. Biological replicates of n=7-9/group. Statistical analyses by one-way ANOVA with Tukey’s test.

**Supplementary Figure 7.**
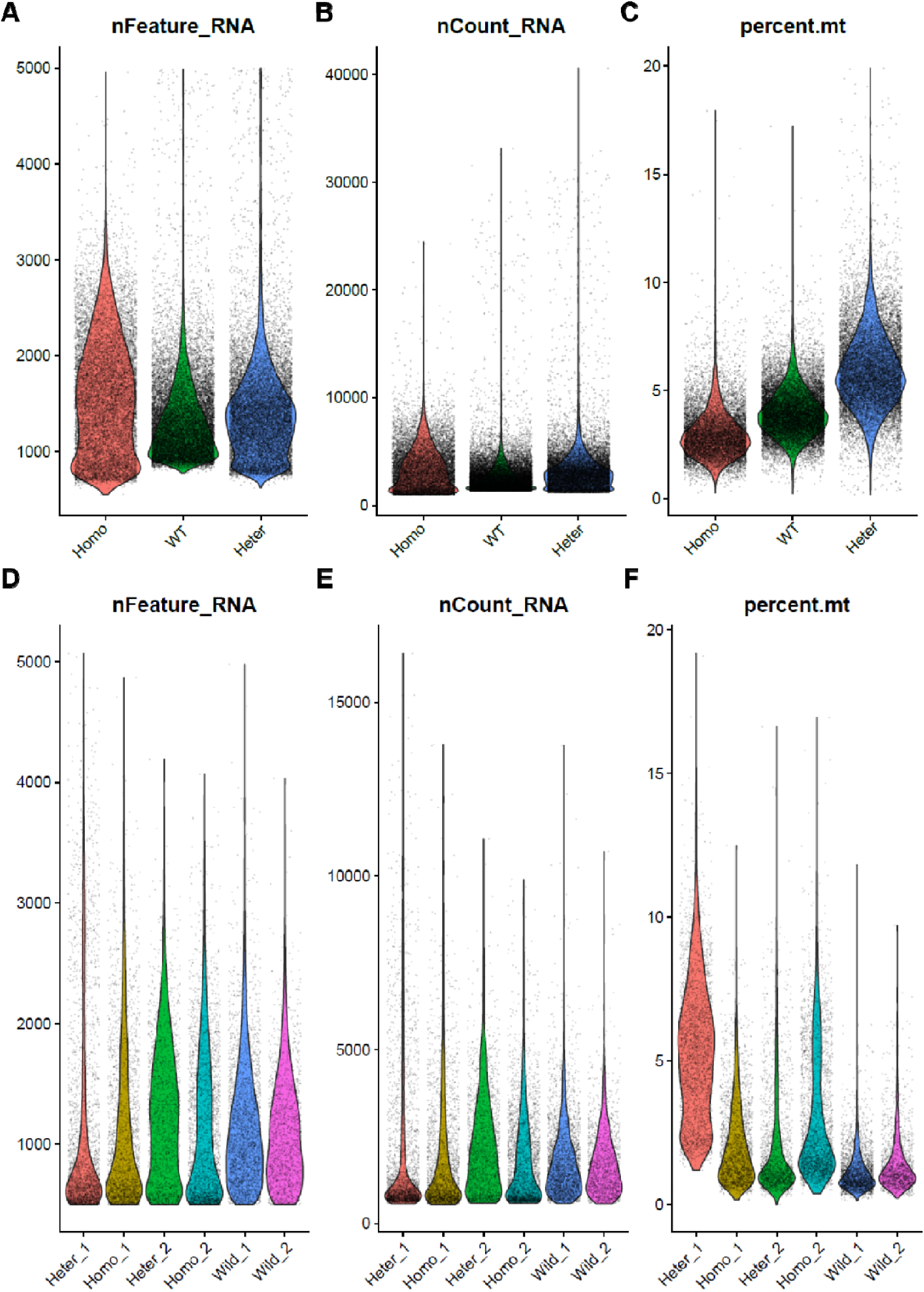
Comprehensive quality control and filtering metrics for single-cell RNA sequencing of superior cervical ganglion (SCG) and atrioventricular nodal (AVN) cells. **(A-C)** Violin plots summarizing the stringent quality-control parameters applied to the SCG scRNA-seq dataset across WT, HET, and HOM rats. The distributions detail the number of detected genes per cell (nFeature_RNA; **A**), the total transcriptomic complexity represented by unique molecular identifiers (nCount_RNA; **B**), and the relative abundance of mitochondrial transcripts (percent.mt; **C**). Sample size = 1 batch. **(D-F)** Violin plots summarizing the stringent quality-control parameters applied to the AVN scRNA-seq dataset across WT, HET, and HOM mutant rats. The distributions detail the number of detected genes per cell (nFeature_RNA; **D**), th total transcriptomic complexity represented by unique molecular identifiers (nCount_RNA; **E**), and the relative abundance of mitochondrial transcripts (percent.mt; **F**). Sample size =2 batch.

**Supplementary Figure 8.**
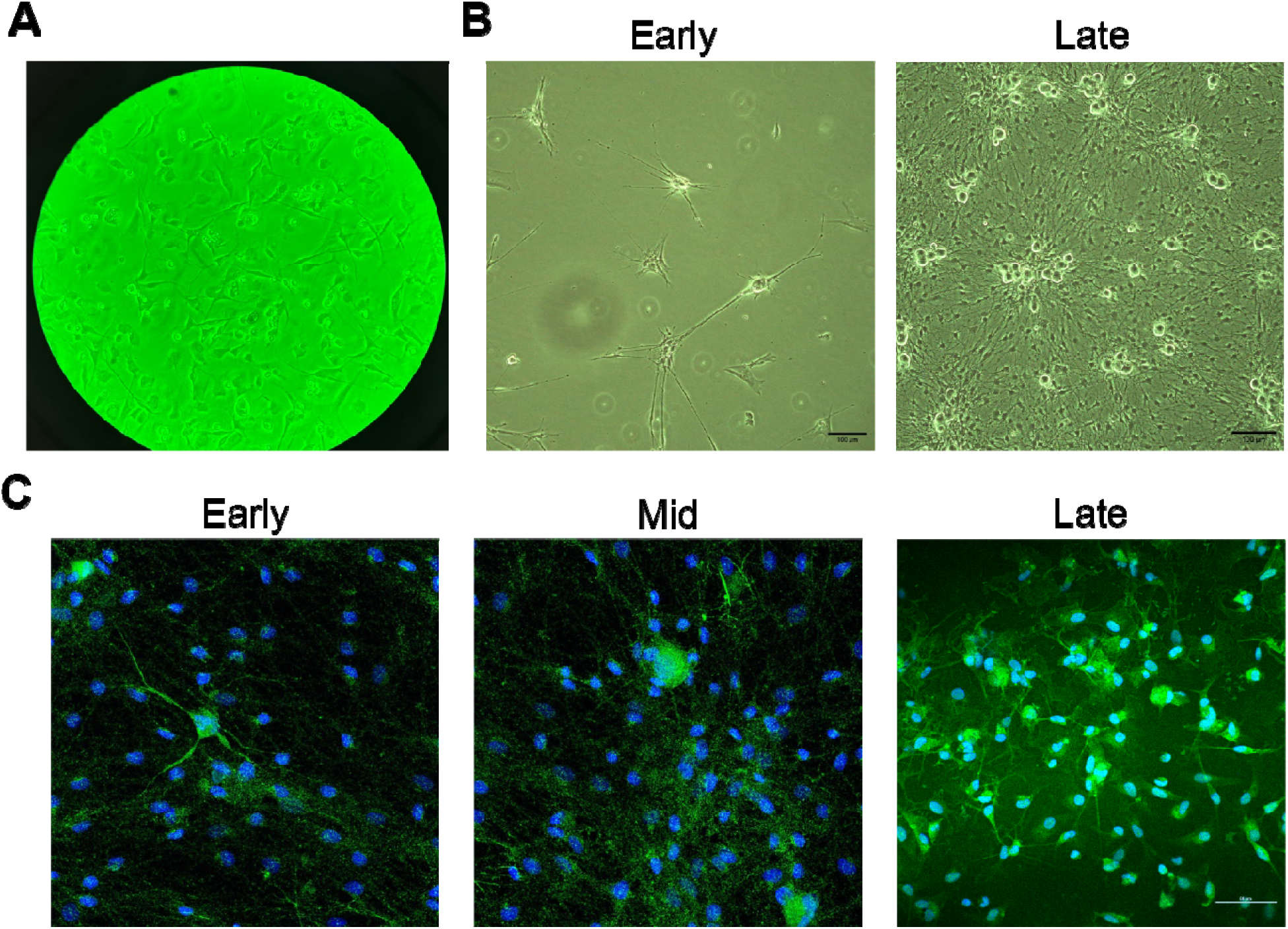
Isolation, culture, and phenotypic validation of primary superior cervical ganglion (SCG) neurons for downstream patch-clamp electrophysiology and subcellular immunofluorescence assays. **(A)** High-resolution phase-contrast imaging illustrating canonical neuronal morphology, exhibiting robust soma integrity, extensive neuritogenesis (neurite outgrowth), and the establishment of complex intercellular networks *in vitro*. **(B)** Representative bright-field micrographs document the successful isolation and early to late *in vitro* adherence of primary sympathetic neurons derived from rat superior cervical ganglia. Scale = 100 μm. **(C)** Confocal immunofluorescence microscopy confirming strict neuronal lineage identity. The primary cultures display uniform immunopositivity for canonical pan-neuronal markers, coupled with DAPI nuclear counterstaining (blue). Scale = 50 μm.

**Supplementary Figure 9.**
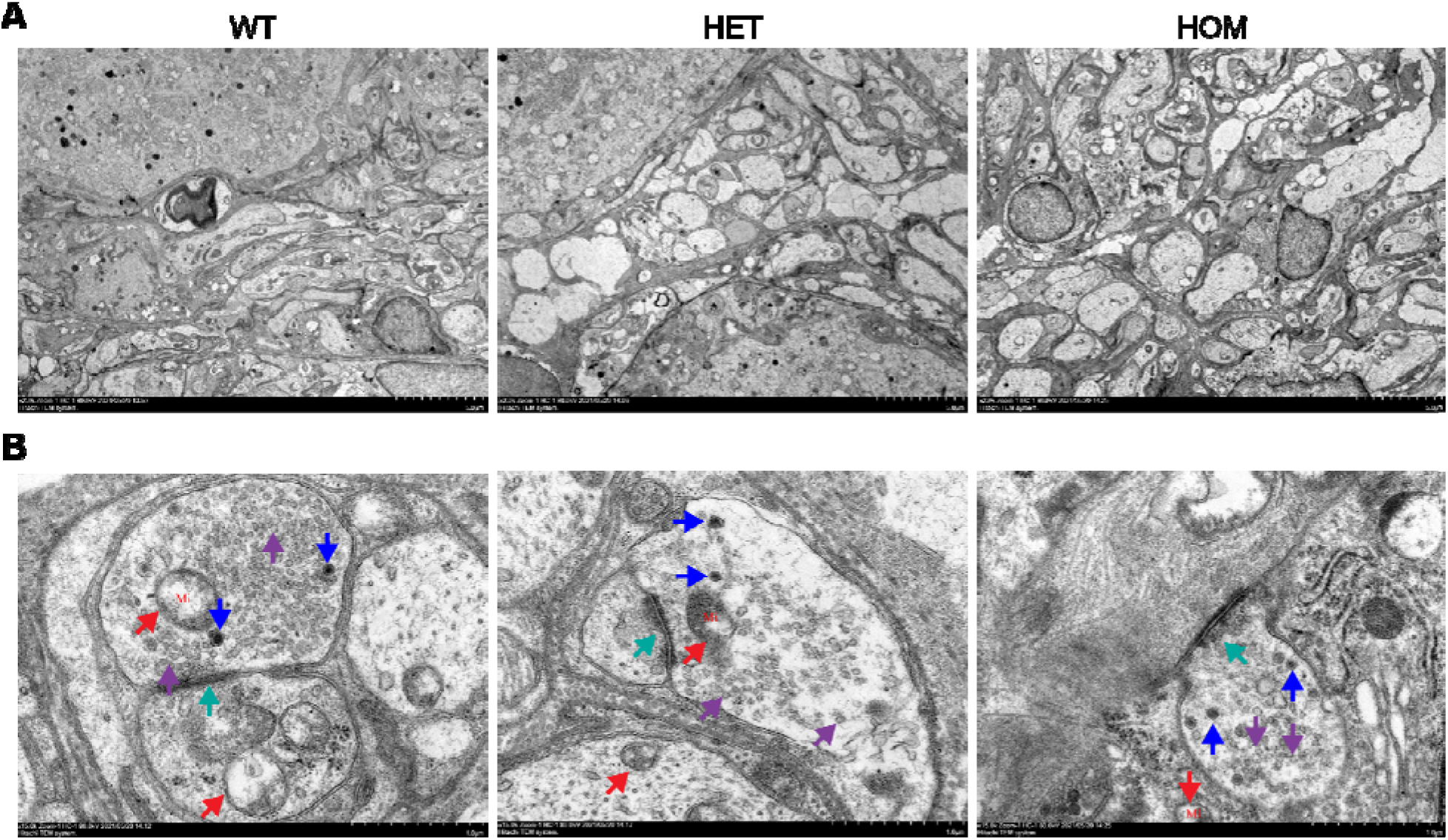
Transmission electron microscopy (TEM) reveals preserved ultrastructural integrity of the SCG tissue across *Cacna1b* genotypes. **(A)** Representative TEM micrographs of native SCG tissues isolated from WT, HET, and HOM rats. **(B)** High-resolution ultrastructural examination demonstrating intact somatic cytoarchitecture in mutants. Key subcellular compartments, including organelle morphology (mitochondria, red arrows; synaptic junction, green arrows) and the density of presynaptic vesicle pools (small and large vesicles, purple and blue arrows), remain conserved across genotypes. This vital morphological evidence conclusively indicates that the arrhythmogenic sympathetic storm is driven by a primary *functional* (biophysical) overdrive of the ganglia, rather than reflecting underlying structural neurodegeneration or inherent organelle toxicity. Scale bar = 50 μm **(A)** and 10 μm **(B)**. Biological replicates of n=6/group.

